# Natural language markers of drug context encoding track neural synchrony and treatment progression in heroin use disorder

**DOI:** 10.1101/2025.09.09.25335113

**Authors:** Sarah G. King, Greg Kronberg, Natalie E. McClain, Zihan Zhang, Ahmet O. Ceceli, Yuefeng Huang, Nelly Alia-Klein, Rita Z. Goldstein

**Author notes:** Corresponding author: Rita Z. Goldstein.

## Abstract

Language transforms subjective internal states into observable behavior, enabling investigation of the neurocognitive dynamics central to psychiatric conditions. Intense emotional and physiological responses to evocative drug-related contexts precipitate craving and drug seeking in substance use disorders, yet sensitive, accessible behavioral markers of the underlying neural dynamics remain elusive. In this fMRI study, we analyzed individuals’ unconstrained verbal recall of their subjective experiences after watching a drug-themed movie, including inpatients in treatment for heroin use disorder (HUD) and healthy controls, with speech recorded outside the scanner under minimal instructions. The semantic context of these accounts, assessed using transformer-based embeddings, effectively predicted both HUD diagnosis and treatment progression, with earlier treatment characterized by heightened self-reference and drug focus but less socially oriented recall. Semantic similarities between individuals reflected synchronized activation patterns in dorsal attention and visual networks during movie viewing, linking motivated attentional processing to later recall. With treatment, most HUD-biased semantic features shifted toward control-like patterns, while drug focus persisted; the semantic-neural link was attenuated though partially remained. These results suggest that spontaneous speech, collected offline, can provide a sensitive behavioral readout of both treatment effects and neural encoding of a complex, personally relevant, drug-related context. offering a scalable approach for monitoring neurocognitive dynamics in real-world settings.

## Introduction

As a complex and uniquely human behavior, natural language offers ecological insights into subjective experiences that elude standard laboratory assessments. By engaging distributed neural systems (Silbert et al., 2014; Hasson et al., 2018) and recruiting higher order functions such as attention, self-regulation, and perspective-taking (Ellis, 2019), spontaneous, unstructured speech may be especially sensitive to cognitive state variations, potentially yielding clinically meaningful indicators of mental health and disease (Cox et al., 2021; Chan et al., 2023; Agurto et al., 2025). However, it is yet unclear whether semantic-neural relationships observed in controlled neuroimaging settings generalize to everyday environments, where language could provide an accessible, behavior-based proxy for neurocognitive mechanisms in psychiatric contexts. In practice, verbal communication remains the primary medium through which psychiatric assessments are made, including for substance use disorders, while also providing the foundation for many therapeutic interventions (psychotherapy, peer support groups, online forums and mental health phone apps); natural language could therefore offer a practical means of probing clinically relevant mental states and behaviors within experimental contexts that are more closely aligned with real-world, subjectively relevant experiences.

Across psychiatric disorders, many core symptoms and their changes with treatment emerge from the subjective experiences of dynamic social and emotional contexts embedded in real-world settings. The inherent heterogeneity of such naturalistic experiences introduces significant ecological complexities that underscore many of the ongoing barriers to translational neuroscience research, including in addiction (Heilig et al., 2016; de Wit et al., 2018). Well-controlled experimental neuroimaging paradigms have yielded important insights into relevant neurocognitive functions and their behavioral correlates (Makowski et al., 2025; Williams and Whitfield Gabrieli, 2025), yet the constrained scope of most laboratory-based assessments potentially limits both their generalizability to more naturalistic contexts and their clinical utility. Identifying scalable and accessible neurocognitive measures of subjective experiences in naturalistic settings could help rectify this missing link by reconciling mechanistic findings with the variable contexts that influence experiential heterogeneity as relevant to symptom expression across psychiatric disorders in the real world.

A core neurocognitive impairment in drug addiction is enhanced salience attribution to drug-related stimuli at the expense of alternative reinforcers; accompanied by impaired behavioral control over drug use, this constellation presents a significant barrier to recovery (Goldstein and Volkow, 2011; Edwards and Koob, 2013; Huang et al., 2024). Studies of brain functioning during cue reactivity demonstrate drug-biased activations in large-scale networks associated with reward, salience, executive, and self-directed functions, while activations in these same systems are reduced in nondrug-related social/emotional, inhibitory control, and decision-making tasks (Zilverstand et al., 2018). These dynamics are thought to contribute to cue-induced craving and relapse, whereby the cognitive appraisal of naturalistic drug-related cues and environments provokes diminished control over further use (Bossert et al., 2013).

According to a recent meta-analysis (Vafaie and Kober, 2022), biological cue reactivity and subjective cue-induced craving are strong indicators of future drug use, further underscoring the importance of resolving neurocognitive dynamics in contexts that are ecologically meaningful.

In our recent study, individuals in inpatient treatment for heroin use disorder (HUD) and healthy controls (CTL) viewed a segment of the addiction-themed movie *Trainspotting* (1996), containing explicit and implicit drug-related content embedded within relevant social and emotional contexts, during fMRI (Kronberg et al., 2024). We observed drug-biased neural response synchronizations between HUD subjects in distributed brain areas associated with reward, salience, and inhibitory control processing, while a partial attenuation of these intersubject correlations with treatment tracked concurrent reductions in cue-induced craving. Here, we extend this framework by evaluating participants’ verbal recall of the same stimulus, testing the hypotheses that linguistic features can (1) capture relevant neurocognitive processes accompanying naturalistic drug context encoding, and (2) elicit behavioral markers of recovery. Using transformer-based large language model embeddings, we first assessed the discrimination between participant groups and treatment time points in latent contextual features of participants’ recall, establishing measurable semantic patterns related to addiction and recovery. Next, we examined interpretable linguistic features, comprising drug-related words as an index of drug-biased attention and pronouns as proxies for self-referential (first-person) and socially oriented (third-person) processing, reasoning that these domains can reflect how one subjectively situates themself within the narrative’s context. Finally, we applied representational similarity analysis to test correspondences between recall semantic similarities and neural synchronizations during movie viewing, assessing unconstrained recall of this salient drug-related context as a scalable behavioral correlate of stimulus encoding.

## Materials and Methods

### Participants

Forty individuals with HUD and 31 CTL participants were recruited through a clinical trial evaluating intervention-related recovery outcomes in opiate addiction (NCT04112186), for which therapy-specific results will be reported separately. Participants completed all movie task procedures twice, during an initial baseline visit (Session 1) and approximately 15 weeks later (Session 2). During the inter-scan interval, HUD participants completed an 8-week group therapy program in addition to their regular inpatient treatment (see below). For the analyses involving fMRI data, one CTL participant was excluded due to a scanner malfunction that resulted in data loss but did not disrupt the movie viewing, and five HUD participants were excluded due to excessive head motion (i.e., 30 CTL/35 HUD participants were included in these analyses). An additional 32 HUD participants and 2 CTL participants who only completed Session 1 were retained as part of a replication dataset for the baseline analyses described below (see Supplementary Methods and Results for details).

All HUD participants were recruited as inpatients enrolled in a treatment program that included prescription opioid medications for opioid use disorder (methadone: n=37, buprenorphine/naloxone: n=3) as well as regular therapy counseling and relapse prevention, trauma, and anger management courses, at a collaborating substance use rehabilitation facility in the New York City metropolitan area (Samaritan Daytop Village, Queens, NY). Age-and gender-matched CTL participants were recruited through advertisements and word-of-mouth in the surrounding community.

Eligibility and participant characteristics were assessed during an initial screening with a comprehensive clinical interview administered by trained research coordinators supervised by clinical psychologists. Diagnostic evaluations included the Mini International Neuropsychiatric Interview, 7^th^ Edition (Sheehan et al., 1998) and the Addition Severity Index, 5^th^ Edition (McLellan et al., 1992). All HUD participants met criteria for opiate use disorder with heroin as their primary substance of choice, and CTL participants did not meet criteria for any current or past substance use disorder. Craving and withdrawal symptoms in HUD participants were assessed with the Heroin Craving Questionnaire (HCQ), a modified version of the Cocaine Craving Questionnaire (Tiffany et al., 1993), and the Short Opiate Withdrawal Scale (SOWS) (Gossop, 1990), and their drug dependence severity was assessed with the Severity of Dependence Scale (SDS) (Gossop et al., 1995). Methadone or buprenorphine use was confirmed through urine toxicology at all screening and study visits. For all participants, current or past nicotine use and alcohol dependence were assessed with the Fagerström Test for Nicotine Dependence (FTND) (Heatherton et al., 1991) and the Short Michigan Alcohol Screening Test (SMAST) (Selzer et al., 1975). A brief medical examination including heart rate, blood pressure, alcohol concentration (with saliva test strips), breath carbon monoxide, and pregnancy testing when applicable (with urine test strips) was administered by trained research coordinators during each visit. Estimated verbal and nonverbal IQ were assessed with the reading subtest of the Wide Range Achievement Test-3 (Jastak and Wilkinson, 1993) and the Matrix Reasoning subtest of the Wechsler Abbreviated Scale of Intelligence (Wechsler, 1999), respectively. Other neuropsychological measures, which were acquired during study visits conducted before participants began the experimental protocol, included assessments for symptoms of depression (Beck’s Depression Inventory, BDI) (Beck et al., 1996) in the past two weeks and anxiety (Beck’s Anxiety Inventory, BAI) (Beck and Steer, 1990) in the past month. Group comparisons on demographic, neuropsychological, and substance use characteristics were performed using two-sample Wilcoxon signed-rank tests for continuous variables and chi-squared tests for categorical variables in R version 6.3.1.

Comorbidities in the HUD participants, which were in partial or sustained remission, included major depression (n=15), anxiety disorders (n=7), post-traumatic stress disorder (n=10), and other substance use disorders (alcohol: n=10; cannabis: n=4; cocaine: n=13; other stimulants: n=2; tranquilizers: n=6). Three CTL participants met criteria for past depression, and one for past anxiety. All participants were otherwise medically healthy.

### Experimental Design and Statistical Analysis

*Movie viewing and recall task*: During both experiment sessions, participants viewed the first 17 minutes and 3 seconds of the movie *Trainspotting* with fMRI acquisition as described in our previous publication (Kronberg et al., 2025). Scans were acquired on a Siemans 3T Skyra (Siemans, Erlangen, Germany) with a 32-channel head coil as part of a 2-hour MRI protocol that included other fMRI tasks (Ceceli et al., 2022, 2024; Huang et al., 2024) and T1-, T2-, and diffusion-weighted scanning (King et al., 2022; Ceceli et al., 2023; Gaudreault et al., 2023, 2024). Details on the scanning procedures and MRI preprocessing steps can be found in our previous publication (Kronberg et al., 2025) and in the Supplementary Methods and Results.

Within 45 minutes of completing the MRI procedures, participants were instructed to recall the movie narrative and describe their experience of watching it while their speech was recorded. A research coordinator gave instructions as follows: “Please verbally recall the movie you just watched, in order, with as much detail as possible. The details are more important than the order. While recounting the movie, please also describe any thoughts, feelings, or emotions you may have experienced. Please tell us what these thoughts were and indicate the specific items in each of the scenes that made you have these feelings, and how strong your emotions were.” Participants performed the recall task uninterrupted, and no minimum or maximum time limit was imposed. That is, unlike paradigms that record speech during scanning or electrophysiological acquisition, here speech was collected outside the MRI environment, with minimal instructions and no task constraints. This allowed us to test whether spontaneous naturalistic recall, recorded in an ecologically valid setting, recovers shared neural encoding of a rich, personally salient movie stimulus. Afterwards, participants completed a quality assurance questionnaire (Supplementary Methods and Results) to assess their memory and comprehension of the movie, indicate whether they had seen it before, and rate their attention and understanding of the movie and the audio and visual quality; these measures did not differ significantly between the groups (Table S1; see Table S2 for the baseline replication sample: here all preprocessing and analytic pipelines were applied identically to the main sample. Replication analyses were exploratory rather than confirmatory).

To investigate potential relationships between the dependent variables and craving, a hallmark symptom of addiction that is a strong predictor of future relapse (Burgess-Hull et al., 2022; Ellis et al., 2022; Vafaie and Kober, 2022), participants were asked to rate their current heroin craving immediately before (baseline craving) and after (task-induced craving) the movie viewing (“Please rate how strong your desire for heroin is on a scale of 0-9”). Following the speech task, participants also rated their feelings of craving (scale of 0-9) related to specific scenes (scene-induced craving), sampled from 35 three-second video clips taken at regular 30-second intervals from the entire movie segment. Session comparisons for these craving and other repeated outcome measures of interest in HUD participants were performed using paired Wilcoxon signed-rank tests (Table 2).

*Speech Preprocessing*: Audio processing and text analysis were performed in Python version 3.10.11. Recordings were transcribed to text using OpenAI’s Whisper automated speech recognition model (Radford et al., 2023). Transcribed texts underwent manual quality assurance processing for accuracy, including removal of interviewer speech and irrelevant participant speech (e.g., asking for clarification on the task instructions). The transcripts were then embedded into 768-dimensional vectors using the all-distilroberta-v1 pretrained model from the SentenceTransformers framework for Bidirectional Encoder Representations from Transformers (BERT) sentence encoding (Reimers and Gurevych, 2019). Briefly, sentence transformers are a deep learning model pretrained on a large corpus of sentence pairs with varying similarities in meaning to capture latent contextual features in short strings of text, which are represented by an output embeddings vector. Embeddings vectors can be compared for closeness using metrics such as the cosine similarity, which measures the or semantic similarity between two vectors in embeddings space (in this case, as the cosine of the angle between the two vectors, such that values closer to-1 indicate greater semantic distance and values closer to 1 indicate greater semantic similarity). Since most of the transcripts exceeded the 512-token limit (mean/standard deviation words per sample: 650/429.8; range: 83-2,479, with approximately one token per 0.75 words), samples were divided into windows of 40 words, equal to approximately half the length of the shortest transcript, to balance preserving local structure while minimizing sparsity. An embeddings vector was extracted for each window, and these were averaged across all the windows from each transcript, resulting in one 768-dimensional vector per sample. Transcripts that were not evenly divisible by the window size were truncated by the length of the remainder.

*Classification Analysis*: To test the decoding of group and session from the 142 transcripts (71 participants with two sessions each), dimensionality reduction of the embeddings vectors was performed with principal component analysis using scikit-learn (Pedregosa et al., 2011). The first 25 principal components, representing a cumulative variance ratio of 75.4%, were selected for classification to avoid overfitting the classifier while retaining a substantial proportion of the overall variability in the data. These steps are in line with previous work suggesting principal component analysis can perform better than other dimensionality reduction techniques in handling contextual embeddings for mental health classification tasks when the number of embeddings dimensions exceeds the sample size (Ganesan et al., 2021). Classifier performance on the principal components, with categorical labels for group and session, was assessed with the scikit-learn function permutation_test_score using a logistic regression classifier with L2 regularization, λ=1, and 10-fold cross validation, with 1000 permutations of shuffled labels used to generate a null accuracy distribution. True accuracy scores above the 95^th^ percentile (p<.05) of the null distribution were considered statistically significant.

*Semantic Similarity Analysis*: To assess the semantic similarity between recall samples, cosine similarity values were computed between every pair of embeddings vectors within the same session using the scikit-learn cosine similarity function (Fig. 2a). For statistical comparisons, the pairwise cosine similarities between each sample and each other sample from the same and other group were averaged separately to compute a mean within-group (CTL-CTL or HUD-HUD) and between-group (HUD-CTL) semantic similarity value for each participant in both sessions.

Additionally, a within-subject/between-sessions similarity value was computed for each participant using the cosine similarity of their embeddings vectors from Sessions 1 and 2. The averaged cosine similarity values were evaluated statistically using a 3 (Comparison: CTL-CTL, HUD-HUD, HUD-CTL) × 2 (Session: 1, 2) mixed ANOVA, and an independent groups t-test was used to compare the within-subject similarity between the groups. Statistical tests were performed in R version 4.4.2 (R Core Team).

*Term Frequency Analysis*: To quantify term frequencies for categories of interest, text samples underwent lemmatization and expansion of contractions using Natural Language Toolkit (NLTK) modules in Python (Wagner, 2010), and word counts were extracted for the whole sample and specific term categories as described in the Supplementary Materials and Methods. Frequencies for each category were calculated by dividing the sum of the individual word counts within a category by the total word count for the sample. Table S3 presents categories and word lists used for this analysis, which included drug-related words and first-person (FP) and third-person (TP) pronouns as well as control categories. Group and session comparisons on term frequencies were analyzed using a 2 (Group: HUD, CTL) × 2 (Session: 1, 2) × 2 (Type: FP, TP) mixed ANOVA for pronouns and 2 (Group) × 2 (Session) mixed ANOVAs for other categories. Statistical tests were performed in R version 4.4.2 (R Core Team).

*Semantic-Neural Inter-subject Representational Similarity Analysis*: To assess the representation of BOLD signal patterns during movie viewing in patterns of semantic expression from movie recall, we performed inter-subject representational similarity analysis (IS-RSA) on pairwise semantic and neural similarity matrices (Kriegeskorte, 2008; Dimsdale-Zucker and Ranganath, 2018; Popal et al., 2019). Semantic similarity matrices consisted of the pairwise cosine similarity values computed between participants’ embeddings vectors (Fig. 2A). Neural similarity matrices consisted of Pearson correlation coefficients (intersubject correlations) computed pairwise on participants’ z-scored BOLD time series data. These were extracted separately for 400 cortical regions of interest (ROIs) from the Schaefer 17-network functional atlas (Yeo et al., 2011) and 16 subcortical ROIs from the Harvard-Oxford subcortical structural atlas (RRID:SCR_001476, provided by FSL v.6.0.3, FMRIB, Oxford, UK), resulting in 416 separate neural similarity matrices in each session. Semantic-neural IS-RSA was performed with Mantel tests, (Mantel, 1967) using the “mantel” package for Python (https://github.com/jwcarr/mantel), on the lower triangle of the semantic similarity matrix with each of the lower triangles of the 416 neural similarity matrices separately. For network-level analyses, these ROIs were consolidated into eight cortical functional networks (control, default mode, dorsal attention, limbic, salience/ventral attention, somatomotor, temporoparietal, and visual) according to their Schaefer Network labels. For each test, this resulted in a Spearman’s rank correlation coefficient with 10,000 permutations of shuffled labels used to generate a null distribution for hypothesis testing. Since only significant positive correlations, indicating co-variability between participants in their verbal recall and BOLD responses, were meaningful to interpretation, hypothesis tests were performed one-tailed on the upper tail.

Comparisons of the Spearman’s correlation coefficients (r_s_) between groups and sessions for each ROI were performed using a Fisher r-to-z transformation: r_s_ values were z-transformed and the absolute difference (Z_diff_) computed as the difference between the z-scores divided by the standard error of the difference. Test statistics were assigned p-values using the Normal Cumulative Distribution Function from SciPy (Virtanen et al., 2020), scipy.stats.norm.cdf, followed by Bonferroni correction (p_corr_) for 416 separate tests for ROI analyses/eight tests for network-level analyses. Regions showing significant (p_corr_<.05) group differences between the correlation coefficients in either session were further examined for significant session differences and r_s_ within the groups.

## Results

### Participant Characteristics

Participant groups did not differ significantly in age, gender, race, or days between sessions, but did differ in education, verbal and nonverbal IQ (CTL>HUD), and in depression and anxiety symptoms and alcohol dependence (HUD>CTL). Since all but one CTL participant and two HUD participants were/were not regularly using nicotine, respectively, no statistical comparison was performed for nicotine use. Statistical tests between the groups for these variables and descriptive statistics for other drug use/addiction severity measures in the HUD group are presented in Table 1 (see Table S4 for the baseline replication sample, which included participants who only had Session 1 data, described in Methods). Correlations were inspected between the language dependent variables (e.g., semantic similarity and term frequency measures) and participant variables that were not relevant to our analyses, but which differed significantly between the groups (e.g., education, verbal and nonverbal IQ, and alcohol dependence); none of these correlations were significant (all p>.05), hence these variables were not considered as covariates.

**Table 1.**
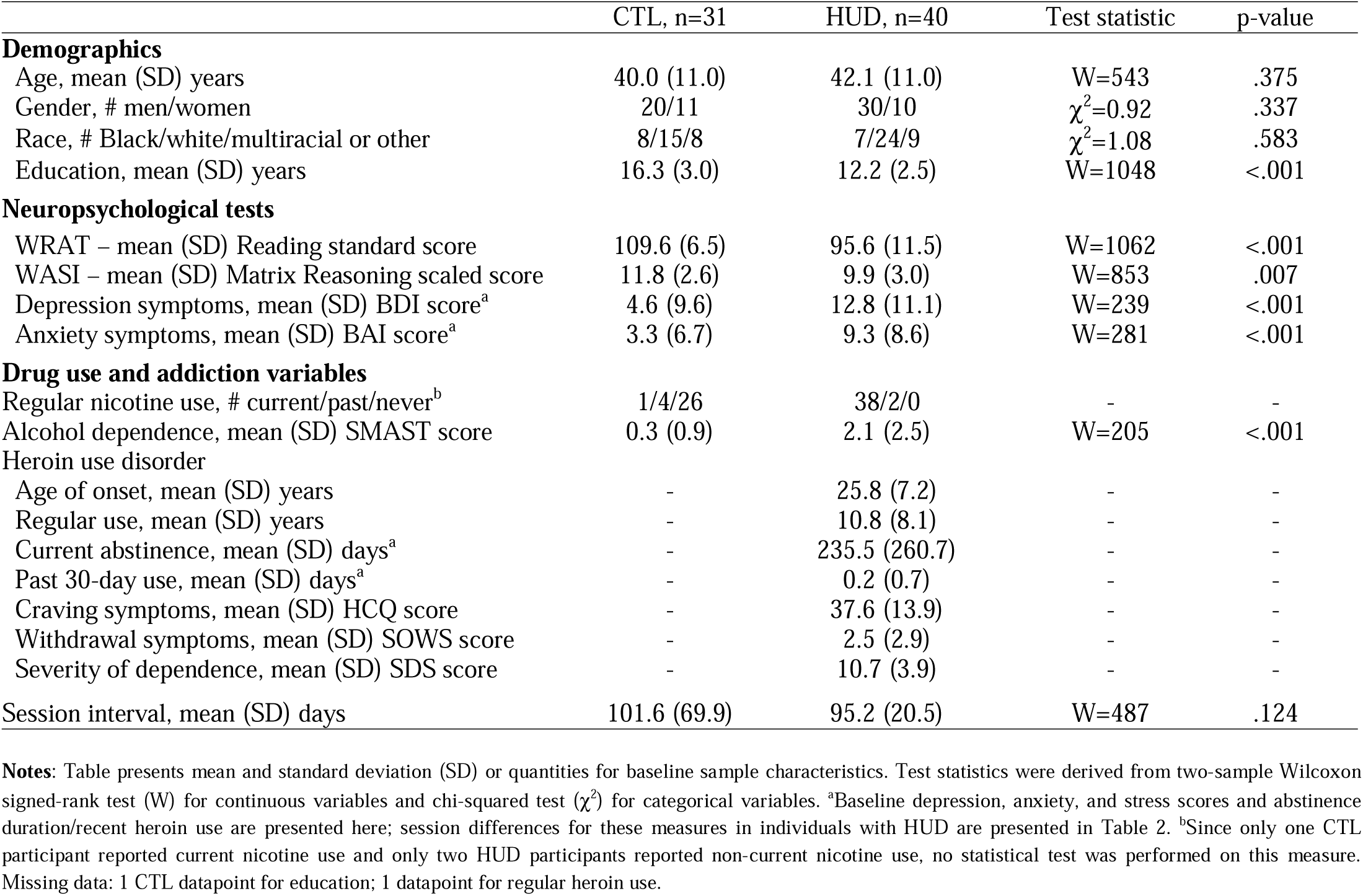
Baseline demographic, neuropsychological, and drug use/addiction measures.

**Table 2.** Repeated neuropsychological and heroin use/craving outcome measures in participants with heroin use disorder.

|  | Session 1 | Session 2 | Test statistic | p-value |
| --- | --- | --- | --- | --- |
| **Neuropsychological tests** |  |  |  |  |
| Depression symptoms, mean (SD) BDI score<sup>a</sup> | 12.8 (11.1) | 11.4 (9.8) | V=414 | .206 |
| Anxiety symptoms, mean (SD) BAI score<sup>a</sup> | 9.3 (8.6) | 10.6 (9.4) | V=274 | .694 |
| **Heroin use and craving variables** |  |  |  |  |
| Current abstinence, mean (SD) days | 235.5 (260.7) | 310.1 (281.8) | V=86 | <.001 |
| Past 30-day use, mean (SD) days | 0.2 (0.7) | 0.7 (2.5) | V=11 | .357 |
| Pre-movie/baseline craving, mean (SD) rating<sup>a</sup> | 2.44 (2.16) | 1.44 (1.82) | V=303 | .006 |
| Post-movie/task-induced craving, mean (SD) rating<sup>a</sup> | 3.00 (2.47) | 2.03 (2.43) | V=289 | .016 |
| Movie craving change, mean (SD) rating difference | 0.56 (1.83) | 0.59 (1.46) | V=151 | .697 |
| Scene-induced craving, mean (SD) rating<sup>a</sup> | 1.19 (1.20) | 0.75 (0.99) | V=408 | .002 |

**Notes:** Table presents mean and standard deviation (SD) for repeated measures in the HUD group. Test statistics were derived from paired Wilcoxon signed-rank test (V). <sup>a</sup>Selected outcome measures were assessed for correlations with test variables of interest. Missing data: 1 datapoint for BDI and BAI; 1 datapoint for each of the movie craving measures.

### Changes in Craving and Severity Measures with Treatment

Comparisons between sessions in the HUD group showed no significant changes over time in depression and anxiety symptoms. As expected for this inpatient population, the mean abstinence duration was significantly longer at Session 2, and there was no significant difference in self-reported past-month heroin use, which had a mean of <1 day at both sessions. Also as expected, there were significant reductions at Session 2 in the pre-movie (baseline) and post-movie (task-induced) craving and the mean scene-induced craving ratings. No significant session difference was observed in the magnitude of the pre-to-post movie change in craving. Statistics for these session comparisons are presented in Table 2.

### Decoding Group and Session from Contextual Embeddings

Classification was tested on the first 25 principal components of 768-dimensional contextual embeddings vectors to inspect discrimination of group and session by semantic features (Fig. 1; see Fig. S2 for the baseline replication sample). A logistic regression classifier performed significantly better than chance (p<.001) at predicting the group and session (4 classes) for all 142 samples, yielding a true accuracy score of 0.49 with a mean null accuracy score of 0.26. In follow-up analyses, classification by group (71 samples/2 classes) yielded a true accuracy of 0.86 with a mean null accuracy of 0.51 (p<.001) at Session 1, and a true accuracy of 0.81 with a mean null accuracy of 0.50 (p<.001) at Session 2. Classification by session (2 classes) yielded a true accuracy of 0.64/mean null accuracy of 0.50 (p=.013) for the 80 samples from the HUD participants. For the 62 samples from CTL participants, classification by session was not significantly significant, yielding a true accuracy of 0.47/mean null accuracy of 0.49 (p=.657), indicating that the linguistic drift over time was specific to treatment-related changes in HUD rather than nonspecific retest or scanner effects.

**Fig. 1.**
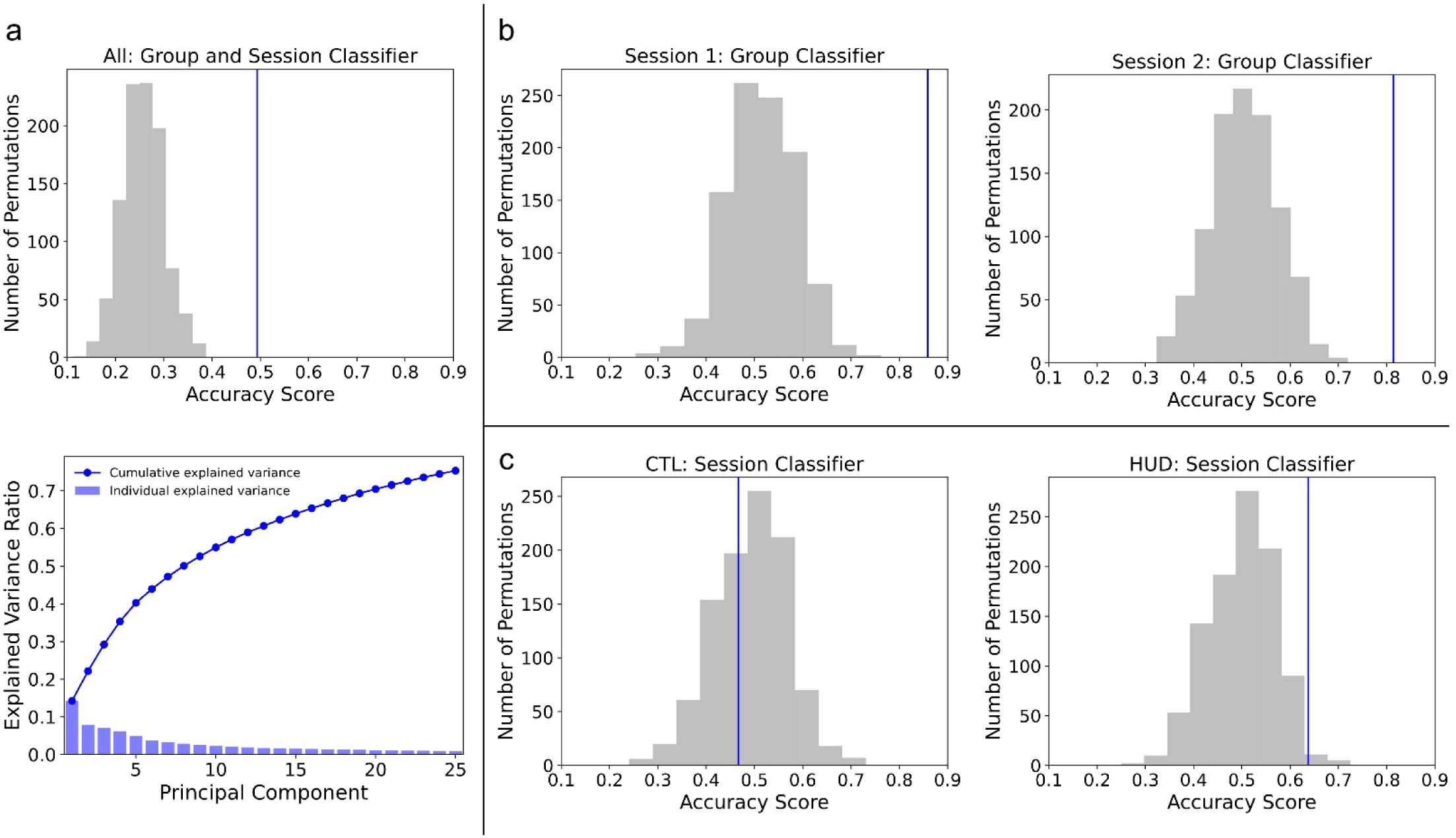
Classifier performance on the first 25 principal components of the embeddings. Principal components explained a cumulative 75.4% of the total variance (variance plot). In the classifier plots, the blue line indicates the true accuracy score, and the gray histogram represents the null accuracy distribution derived from 1,000 permutations of shuffled labels. Classification accuracy was significantly better than chance in decoding a: both the group and session labels from all samples (0.49), b: group labels from Session 1 (0.86) and Session 2 (0.81) samples, and c: session labels from HUD (0.64), but not CTL (0.47), samples.

**Fig. 2.**
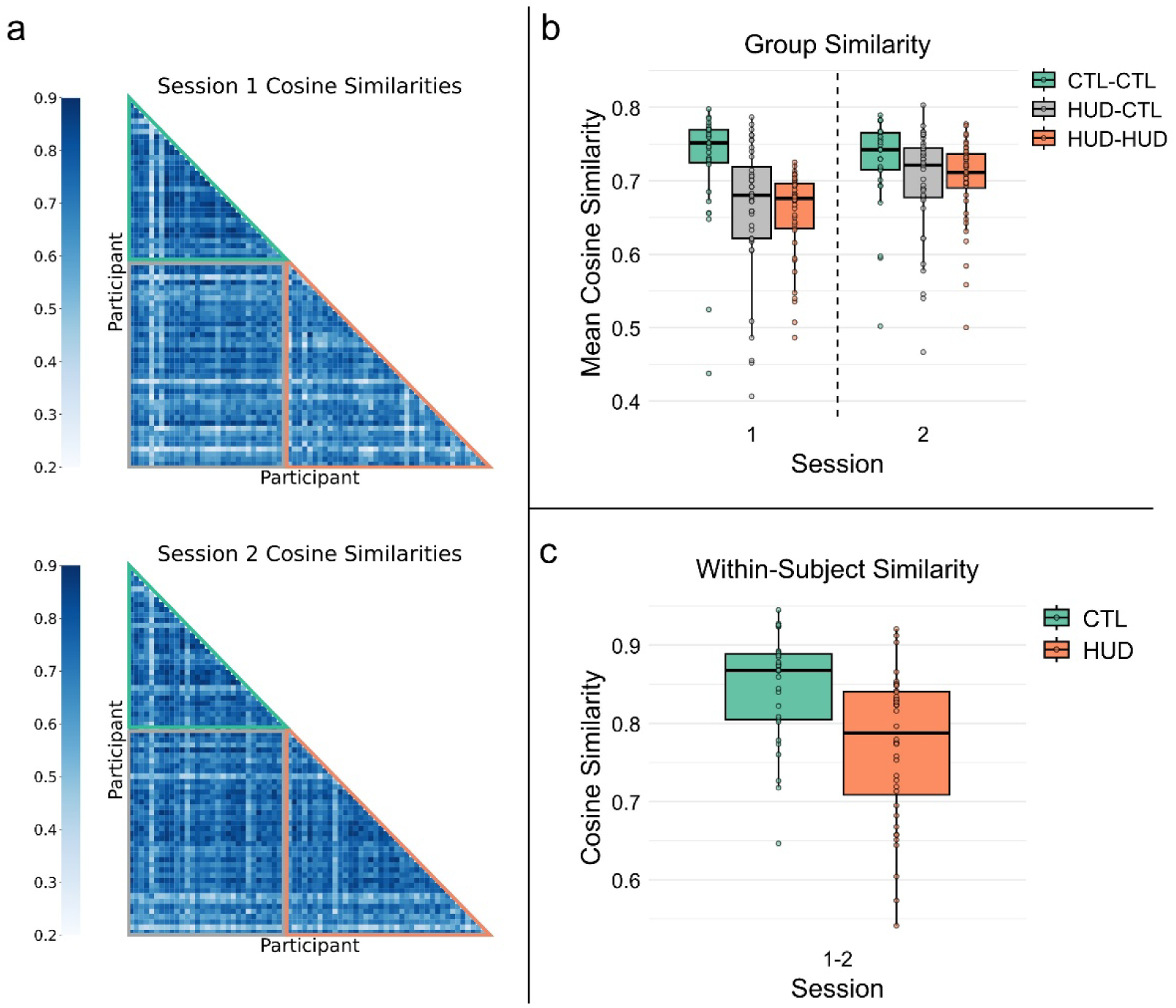
Semantic similarity derived from the embeddings vectors of the speech samples. a: Lower triangle of the pairwise similarity matrices from Session 1 and Session 2. Rows and columns represent the 71 participants, sorted by group (green upper border: CTL; orange lower border: HUD), with pairwise cosine similarity values indicated by the scale bar. b. Mean pairwise CTL-CTL similarity was significantly higher than HUD-CTL and HUD-HUD similarities at Session 1, with no significant differences at Session 2, and both HUD-CTL and HUD-HUD similarities increased between the sessions. c: CTL within-subject similarity was significantly higher than HUD, indicating greater change in HUD participants’ speech between the two sessions. In the boxplots, the center line indicates the median, the box indicates the interquartile range, and whiskers extend to the most extreme points within 1.5x the interquartile range above and below the box.

### Semantic Similarity by Group and Session

Semantic similarity of the embeddings was assessed between subjects within (CTL-CTL, HUD-HUD) and between (HUD-CTL) the groups (Fig. 2b; see Fig. S3 for the baseline replication sample), and within subjects (i.e., between the two sessions for the same subject) (Fig. 2c). A mixed ANOVA for Comparison (CTL-CTL, HUD-HUD, HUD-CTL) and Session (1, 2) revealed significant main effects of both Comparison (F_1,78_=8.13, p=.006) and Session (F_1,78_=12.79, p<.001), with a significant 2-way interaction effect (F_2,78_=5.28, p=.025). Follow-up 1-way ANOVAs showed a significant main effect of Comparison in Session 1 (F_2,108_=8.21, p<.001), with the Tukey HSD post hoc test showing CTL-CTL>HUD-HUD (p<.001), CTL-CTL>HUD-CTL (p=.003), and HUD-HUD=HUD-CTL (p=.876); there was no significant effect at Session 2 (F_2,108_=1.70, p=.189). Paired t-tests for the effect of Session showed Session 1=2 for CTL-CTL (t_30_=0.09, p=.927) and Session 2>1 for HUD-HUD (t_39=_-4.19, p<.001) and HUD-CTL (t_39_=-2.74, p=.009). For the within-subject/between-sessions cosine similarity, an independent groups t-test showed CTL>HUD (t_69_=3.82, p<.001). Thus, HUD intersubject semantic similarities increased from Session 1 to Session 2, approaching CTL-CTL similarity levels, consistent with treatment-related normalization of semantic structure.

### Group and Session Comparisons on Drug Bias and Self vs. Other Reference

To better understand the observed patterns of group and session effects in the embeddings, we examined semantic features that were hypothesized to occur more frequently in the HUD group’s recall. These included drug-biased speech and self-reference vs. socially-directed speech, which were approximated as the frequencies of drug terms and first-person (FP) vs. third-person (TP) pronouns, respectively, in the sample (Fig. 3; see Fig. S4 for the baseline replication sample). For drug term frequencies, a mixed ANOVA examining Group (CTL, HUD) and Session (1, 2) revealed a significant main effect of Group (HUD>CTL, F_1,69_=5.84, p =.018), and no other main or interaction effects (F≤0.44, p≥.510).

**Fig. 3.**
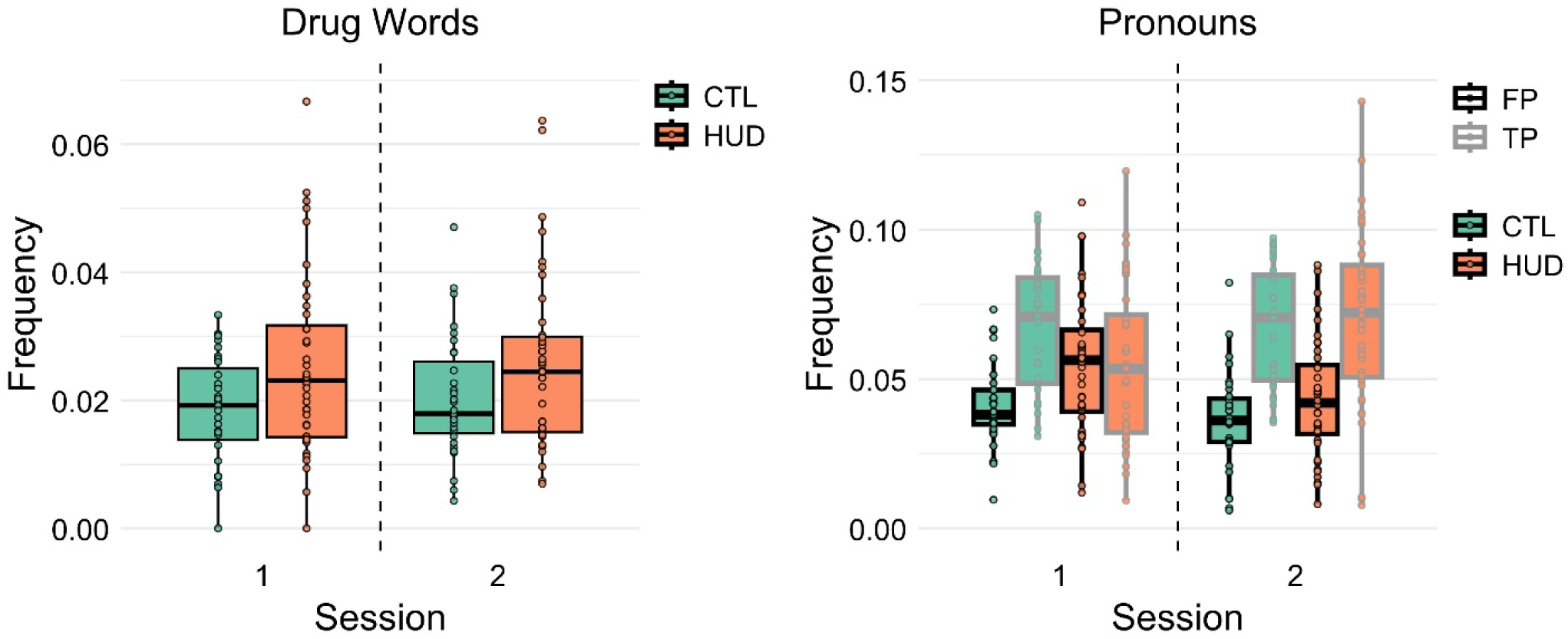
Term frequency analyses for selected categories of interest. Across both sessions, the HUD group referred to drugs significantly more than the CTL group, with no significant change between the sessions in either group. At Session 1, HUD participants used significantly more FP and less TP pronouns than CTL, with no significant differences between the groups at Session 2. Between the sessions, HUD participants showed decreased use of FP and increased use of TP pronouns, while CTL participants showed no significant changes in pronoun use. In the boxplots, the center line indicates the median, the box indicates the interquartile range, and whiskers extend to the most extreme points within 1.5x the interquartile range above and below the box.

The mixed ANOVA examining pronoun term frequencies by Group (CTL, HUD), Session (1, 2), and Type (FP, TP) revealed a significant main effect of Type (F_1,69_=25.86, p<.001), a significant 2-way interaction effect of Session × Type (F_1,69_=13.65, p<.001), and a significant 3-way interaction effect of Group × Session × Type (F_1,69_=6.35, p=.014); there were no other main or interaction effects (F≤3.85, p≥.054). For Session 1, the follow-up ANOVA for the Group × Type effect in the 3-way interaction showed a significant main effect of Type (F_1,138_=10.08, p=.002), no significant main effect of Group (F<0.01, p=.991), and a significant 2-way interaction effect (F_1,138_=13.54, p<.001), with independent groups t-tests showing HUD>CTL for FP (t_69_=-3.12, p=.003) and CTL>HUD for TP (t_69_=2.43, p=.018). There was a significant main effect of Type (F_1,138_=56.44, p<.001) at Session 2, with a paired t-test showing TP>FP (t_70_=-6.17, p<.001), and no other significant main or interaction effects (F≤1.80, p≥.180). A repeated-measures ANOVA for the Session × Type effect in the 3-way interaction showed, for HUD, a significant main effect of Type (F_1,39_=5.08, p=.030) and a significant 2-way interaction effect (F_1,39_=14.45, p<.001), with paired t-tests showing Session 1>2 for FP (t_39_=3.00, p=.005) and Session 2>1 for TP (t_39_=-3.31, p=.002). The same test for CTL showed a significant effect of Type, TP>FP (F_1,30_=34.64, p<.001). Paired t-tests comparing FP and TP frequencies showed, in HUD, no significant difference between pronoun types at Session 1 (t_39_=0.03, p=.972), and TP>FP at Session 2 (t_39_=-3.82, p<.001). The same tests in CTL showed TP>FP at both Session 1 (t_30_=-5.00, p<.001) and Session 2 (t_30_=-5.47, p<.001).

In summary, HUD participants used more FP pronouns and fewer TP pronouns at baseline, indicating heightened self-focus and reduced socially oriented processing. By Session 2, these differences had largely normalized. Unlike pronoun use, drug-related word frequency did not significantly decrease with treatment, suggesting a persistent attentional bias toward drug cues despite broader semantic normalization.

Additional term frequency analyses for control categories, which included place words and non-pronoun stop words, showed no significant effects (Fig. S1).

### Correlations with Outcome Measures

Language variables of interest (mean within-and between-group semantic similarity and drug word, FP pronoun, and TP pronoun frequencies) were inspected for correlations with craving measures that showed significant decreases at Session 2 (baseline, task-induced, and scene-induced craving) as well as depression and anxiety symptom scores in the HUD group.

Spearman’s correlation coefficients for these variables were assessed with their Session 1 measures and deltas (Session 2-Session 1). These tests yielded no significant results for the baseline (|r_s_|≤0.28, p≥.086), task-induced (|r_s_|≤0.25, p≥.124), or scene-induced (|r_s_|≤0.14, p≥.388) craving measures, or for depression (|r_s_|≤0.25, p≥.133) and anxiety (|r_s_|≤0.29, p≥.069) symptoms.

### Speech Representational Similarity to fMRI

To assess the extent to which variabilities in recall reflect neural activation patterns during movie viewing, we performed semantic-neural inter-subject representational similarity analyses (IS-RSA) using the speech cosine similarities and intersubject correlations (ISCs) in fMRI time series (Fig. 4a). First, we inspected ISCs in eight cortical functional networks to assess group similarities in neural activity during movie viewing among HUD and CTL participants (Supplementary Materials and Methods). Compared to CTLs, group-level intersubject correlations in the HUD group were reduced in dorsal attention, visual, and default mode networks at Session 1 (Fig. S5), paralleling the observed semantic variability; HUD ISCs in dorsal attention and visual networks also increased significantly at Session 2 (see Supplementary Materials and Methods for details).

**Fig. 4.**
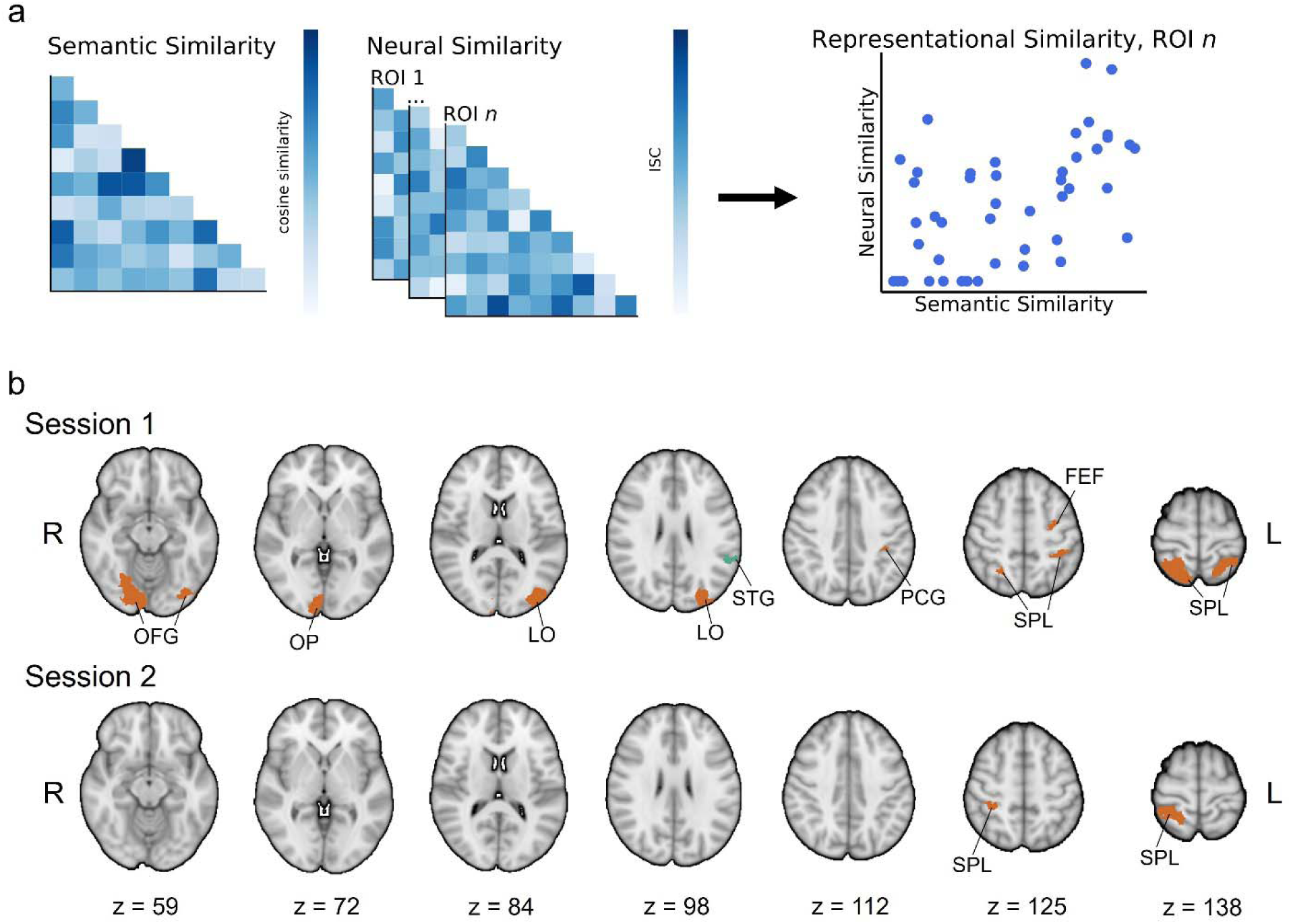
Summary of the RSA comparisons by group and session. a: Per session, correlations between one semantic similarity matrix (also see Fig. 2a) and separate neural similarity matrices per region of interest (ROI) were computed using Mantel tests with Spearman’s correlation coefficients. b: Anatomical distribution of the RSA results. Regions in which the Spearman’s correlation was significantly higher in the HUD vs. CTL (orange) or CTL vs. HUD group (green), and which also showed a significant positive correlation in the HUD or CTL group (see Table 3), are shown here. OFG: occipital fusiform gyrus; OP: occipital pole; LO: lateral occipital cortex; STG: superior temporal gyrus; PCG: postcentral gyrus; SPL: superior parietal lobule; FEF: frontal eye field.

**Table 3.** Statistics for semantic-neural representational similarity analysis.

| Structure (Schaefer Atlas #) | Correlation Comparison, $Z_{\text{diff}}$ ($p_{\text{corr}}$) | | | | Session 1, $r_s$ (p) | | Session 2, $r_s$ (p) | |
| --- | --- | --- | --- | --- | --- | --- | --- | --- |
|  | Group: Session 1 | Group: Session 2 | Session: HUD | Session: CTL | CTL | HUD | CTL | HUI |
| **HUD>CTL** |  |  |  |  |  |  |  |  |
| Left Occipital Fusiform Gyrus (3)<sup>1</sup> | **5.30 (<.001)** | -0.58 (>.999) | **5.83 (<.001)** | - | -0.09 (.755) | **0.24 (.008)** | -0.06 (.705) | -0.09 (.8 |
| Left Lateral Occipital Cortex (10)<sup>1</sup> | **6.26 (<.001)** | **4.78 (.001)** | 2.18 (>.999) | - | -0.16 (.868) | **0.24 (.015)** | -0.19 (.924) | 0.11 (.1 |
| Left Lateral Occipital Cortex (12)<sup>1</sup> | **5.89 (<.001)** | 2.00 (>.999) | **5.29 (<.001)** | - | -0.05 (.615) | **0.32 (.004)** | -0.11 (.797) | 0.02 (.4 |
| Left Lateral Occipital Cortex (65)<sup>1</sup> | **5.73 (<.001)** | **3.87 (.046)** | 3.37 (.308) | - | -0.06 (.650) | **0.30 (.004)** | -0.14 (.856) | 0.11 (.1 |
| Left Postcentral Gyrus (77)<sup>1</sup> | **5.26 (<.001)** | 3.43 (.254) | 1.42 (>.999) | - | -0.14 (.902) | **0.19 (.009)** | -0.11 (.864) | 0.11 (.0 |
| Left Superior Parietal Lobule (78)<sup>1</sup> | **5.52 (<.001)** | **3.94 (.033)** | 1.05 (>.999) | - | -0.16 (.901) | **0.18 (.007)** | -0.13 (.853) | 0.12 (.0 |
| Left Superior Parietal Lobule (80)<sup>1</sup> | **5.97 (<.001)** | **4.34 (.006)** | 1.83 (>.999) | - | -0.17 (.904) | **0.21 (.015)** | -0.17 (.924) | 0.10 (.1 |
| Left Frontal Eye Field (84)<sup>1</sup> | **3.89 (.041)** | 0.05 (>.999) | **4.82 (<.001)** | - | -0.02 (.571) | **0.22 (.006)** | -0.06 (.695) | -0.05 (.8 |
| Right Occipital Fusiform Gyrus (203)<sup>1</sup> | **4.31 (.007)** | 2.68 (>.999) | 3.48 (.206) | - | -0.07 (.682) | **0.20 (.043)** | -0.17 (.893) | >-0.01 (.0 |
| Right Occipital Fusiform Gyrus (204)<sup>1</sup> | **4.34 (.006)** | 2.45 (>.999) | 3.50 (.190) | - | -0.04 (.603) | **0.23 (.006)** | -0.12 (.836) | 0.03 (.6 |
| Right Occipital Pole (207)<sup>1</sup> | **4.62 (.002)** | 1.97 (>.999) | 3.65 (.107) | - | -0.07 (.694) | **0.22 (.002)** | -0.12 (.831) | >-0.01 (.0 |
| Right Lateral Occipital Cortex (210) | **4.24 (.009)** | 2.56 (>.999) | 1.28 (>.999) | - | -0.08 (.720) | 0.18 (.054) | -0.05 (.640) | 0.11 (.1 |
| Right Occipital Pole (211) | **5.06 (<.001)** | 3.28 (.430) | 0.12 (>.999) | - | -0.18 (.924) | 0.14 (.074) | -0.07 (.725) | 0.13 (.0 |
| Right Lateral Occipital Cortex (212) | **4.41 (.004)** | **4.31 (.006)** | 0.54 (>.999) | - | -0.12 (.799) | 0.15 (.078) | -0.15 (.859) | 0.12 (.1 |
| Right Superior Parietal Lobule (271)<sup>1</sup> | **5.22 (<.001)** | 2.29 (>.999) | 3.29 (.421) | - | -0.05 (.638) | **0.27 (.003)** | -0.06 (.650) | 0.02 (.2 |
| Right Superior Parietal Lobule (272)<sup>1</sup> | **3.87 (.045)** | 2.50 (>.999) | 2.97 (>.999) | - | -0.05 (.656) | **0.19 (.025)** | -0.14 (.870) | 0.02 (.4 |
| Right Superior Parietal Lobule (278)<sup>2</sup> | 2.62 (>.999) | **5.24 (<.001)** | -1.27 (>.999) | - | -0.04 (.601) | 0.12 (.064) | -0.13 (.870) | **0.19 (.0** |
| Right Superior Parietal Lobule (280)<sup>1,2</sup> | **5.27 (<.001)** | **5.61 (<.001)** | 0.04 (>.999) | - | -0.12 (.813) | **0.21 (.015)** | -0.14 (.879) | **0.21 (.0** |
| **CTL>HUD** |  |  |  |  |  |  |  |  |
| Left Supramarginal Gyrus (87)<sup>1</sup> | **-4.49 (.003)** | -1.34 (>.999) | - | 2.75 (>.999) | **0.29 (.003)** | 0.01 (.387) | 0.11 (.119) | 0.03 (.3 |

**Notes:** Table presents statistics for structures that met criteria for significance in representational similarity analysis of recall semantic similarity and BC ISC during movie viewing in the replication sample. Comparisons of the correlation coefficients between groups/within each session, and between sessions within the groups, were performed using Fisher Z testing ( $Z_{\text{diff}}$ ) with Bonferroni-corrected p values, shown in the Correlation Comparison column. Spearman $r$ ( $r_s$ ) and uncorrected p-values derived from Mantel tests are shown for each group. Bold values indicate statistically significant tests ( $p < .05$ ). Regions that showed a significant group difference in the correlation coefficient and a significant RSA correlation within a group are denoted <sup>1</sup> for Session 1 and <sup>2</sup> for Session 2.

IS-RSAs were carried out in the same eight networks, with BOLD ISCs in dorsal attention and visual networks showing significantly greater representational similarity to movie recall embeddings in HUD compared to CTL at Session 1. No significant IS-RSA group differences were observed at Session 2, and the visual network representational similarity decreased between sessions in HUD (Table S5; see Table S6 for the baseline replication sample).

We then inspected similar comparisons in 416 individual regions of interest (ROIs), including 400 cortical parcellations of the same eight functional networks and 16 subcortical areas, to assess the specificity of the IS-RSA results. This yielded 14 ROIs in which the representational similarity was significantly higher in the HUD vs. CTL group (p_corr_<.05) at Session 1, driven by significant positive correlations in HUD. Consistent with the network-level analyses, these significant ROIs encompassed cortical areas associated with the dorsal attention and visual networks, including the left dorsolateral prefrontal cortex/frontal eye field, bilateral superior parietal lobule, left postcentral gyrus, left medial and lateral occipital cortex and right occipital pole, and bilateral occipital fusiform gyrus. Significant CTL>HUD representational similarity was observed in one ROI in the left supramarginal gyrus. At Session 2, two ROIs in the right superior parietal lobule showed significant HUD>CTL representational similarity. The IS-RSA correlations in three ROIs in the left dorsolateral prefrontal cortex/frontal eye field, left lateral occipital cortex, and left occipital fusiform gyrus decreased significantly in the HUD group from Session 1 to 2. Results from the ROI analyses are presented in Table 3 and Fig. 4b (see Table S7 and Fig. S6 for the baseline replication sample).

In summary, spontaneous speech recorded offline and outside the imaging environment significantly recapitulated neural synchronization patterns elicited by the movie, demonstrating that shared semantic structure in recall mirrors shared neural encoding. Specifically, these results show that semantic organization of spontaneous recall selectively reflected activity in dorsal attention and visual regions, highlighting top-down attentional encoding in shaping subsequent verbal recall.

## Discussion

Naturalistic language offers a powerful window into how the brain encodes and organizes experience in real-world contexts. The present findings highlight the potential of naturalistic language to reveal stable individual differences in core neurocognitive functions, showing how unconstrained verbal recall can reflect semantic and attentional processing demands from a complex stimulus. The alignment between semantic variability and shared neural activation patterns in dorsal attention and visual networks suggests that spontaneous speech can capture how motivated attention shapes naturalistic perception, extending evidence that attention reorganizes distributed population codes to prioritize behaviorally relevant information (Nastase et al., 2017). Critically, these links between speech and neural encoding were observed using spontaneous recall recorded outside the MRI environment, under minimal task constraints, indicating that scalable, low-burden speech measures can index shared neurocognitive responses to a rich, personally salient and dynamic stimulus without simultaneous speech acquisition.

These results therefore contribute to a growing theoretical model in which language serves not only as an expressive output, but also as an interpretable trace of underlying neurocognitive dynamics.

This framework carries broad implications for psychiatry, where many symptoms emerge in naturalistic, emotionally salient environments that are rarely captured in controlled laboratory settings. Quantifiable linguistic features have already demonstrated utility for diagnostic classification and outcome prediction across disorders (Malgaroli et al., 2023), yet most prior work has relied on clinical notes or passive text sources rather than experimentally elicited speech. By linking semantic patterns to neural encoding of a shared stimulus, the current study provides mechanistic support for using naturalistic speech as an accessible behavioral proxy for neurocognitive states. Such measures may help bridge the long-standing gap between neuroimaging findings and the real-world contexts that shape symptom expression (Makowski et al., 2025), offering a low-burden approach for monitoring neurocognitive processes across psychiatric conditions.

For addiction specifically, this study identifies linguistic markers that reflect hallmark cognitive disruptions, including drug-biased attention, heightened self-focus, and less socially oriented processing. At baseline, these features were more pronounced among individuals with HUD, partially normalizing with treatment. This variability likely reflects the diverse personal histories and environmental contexts that shape drug-motivated attention, which in turn influence both neural encoding and later recall. Consistent with models of dysregulated salience attribution (Goldstein and Volkow, 2011), drug-related language may index biased attentional allocation toward drug cues, whereas self-referential language may reflect heightened personal identification with drug-themed content. Importantly, semantic-neural correspondences were strongest in dorsal attention and visual regions, including superior parietal lobule, frontal eye field, and occipital cortex, suggesting that drug-motivated attention sculpts sensory encoding of naturalistic scenes and shapes the semantic representations recovered during recall. These results complement cue reactivity research, which typically focuses on valuation circuits, by highlighting attentional and sensory dynamics that may persist over longer timescales (Patai et al., 2012; Nobre and Stokes, 2019).

Despite encouraging findings, this study has limitations. We did not identify associations between linguistic features and clinical measures such as craving, depression, or anxiety, potentially due to the relatively short (∼3 month) follow-up interval; it is also possible that language captures more implicit processes than clinical measures based on self-evaluative, declarative processes. Longer-term longitudinal designs, with denser sampling across treatment and post-discharge, will be needed to establish the predictive validity of these linguistic markers. In addition, the present findings were obtained in a specific drug-related narrative context; future studies should assess generalizability to neutral or other personally relevant contexts and associations with drug use and/or relapse risk in real-world environments.

To conclude, we show that unstructured verbal recall provides a scalable and ecologically relevant assay of neural encoding in addiction. By capturing systematic links between semantic patterns, treatment progression, and attentional encoding, this approach broadens the methodological toolkit for basic research on naturalistic cognition and offers clinically relevant opportunities for digital phenotyping, treatment monitoring, and outcome prediction in addiction and beyond.

## Supporting information

Supplementary Methods and Results

## Data Availability

All data produced in the present study are available upon reasonable request to the authors

