## Supplementary Methods and Results for "Natural language markers of drug context encoding track neural synchrony and treatment progression in heroin use disorder"

MRI acquisition and preprocessing

The following descriptions of image acquisition and preprocessing steps were adapted from our previous publication (Kronberg et al., 2025). Participants watched the movie, which was displayed on a screen outside the bore of the scanner, through a mirror mounted on the head coil. MRI compatible in-ear headphones were used for audio. During the movie viewing, blood oxygen level-dependent (BOLD) signals were recorded with a T2*-weighted single shot multi-band gradient-echo EPI sequence (time to echo/repetition time=35/1000 ms, 2.1 mm isotropic voxel resolution, 70 whole-brain axial slices, 206 × 181 mm field of view (FOV), 96 × 84 matrix size, 60° flip angle, Controlled Aliasing in Parallel Imaging Results in Higher Acceleration/CAIPIRINHA phase-encoding shift=FOV/3, ~2 kHz/pixel bandwidth with ramp sampling, 0.68 ms echo spacing/57.1 ms echo train length). T1-weighted structural scans were acquired with a 3D Magnetization-Prepared Rapid Gradient-Echo/MPRAGE sequence (time to echo/repetition time/inversion time=2.07/2400/1000 ms, 0.8 mm isotropic voxel resolution, 256 × 256 × 179 mm3 FOV, 8° flip angle with binomial (1, -1) fat saturation, 240 Hz/pixel bandwidth, 7.6 ms echo spacing, in-plane acceleration (generalized autocalibrating partially parallel acquisitions/GRAPPA) factor of 2).

Preprocessing of T1-weighted structural scans was performed with fMRIprep version 20.2.1 (Esteban et al., 2019). Scans were individually corrected for intensity non-uniformity with N4BiasFieldCorrection (Tustison et al., 2010), distributed by ANTs 2.3.3 (Avants et al., 2008), to be used as T1-weighted structural reference images for fMRI preprocessing. The reference image was skull-stripped with a Nipype implementation of the antsBrainExtraction.sh workflow from ANTs using OASIS30ANTs as the target template. Brain tissue segmentation of CSF, white matter and grey matter was performed on the brain-extracted T1-weighted image using FAST (FSL 5.0.9, RRID: SCR_002823) (Zhang et al., 2001). Volume-based spatial normalization to a standard space (MNI152NLin2009cAsym) was performed through non-linear registration with antsRegistration (ANTs 2.3.3) using brain-extracted versions of both the T1-weighted reference and template. The ICBM 152 Nonlinear Asymmetrical template version 2009c (RRID:SCR_008796; TemplateFlow ID: MNI152NLin2009cAsym) (Fonov et al., 2011) was used for spatial normalization.

Functional image preprocessing was also performed with fMRIprep version 20.2.1. First, a reference volume and its skull-stripped version were generated by aligning and averaging a single-band reference. A B0 non-uniformity map (or fieldmap) was estimated based on two EPI references with opposing phase- encoding directions, with AFNI’s 3dQwarp (Cox and Hyde, 1997). Based on the estimated susceptibility distortion, a corrected EPI reference was calculated for a more accurate co-registration with the T1-weighted structural reference. The BOLD reference was then co-registered to the T1-weighted reference using FLIRT (FSL 5.0.9) (Jenkinson and Smith, 2001) with the boundary-based registration cost-function (Greve and Fischl, 2009). Co-registration was configured with nine degrees of freedom to account for distortions remaining in the BOLD reference. Head-motion parameters with respect to the BOLD reference (transformation matrices and six corresponding rotation and translation parameters) were estimated before any spatiotemporal filtering using MCFLIRT (FSL 5.0.9) (Jenkinson et al., 2002). The BOLD time-series were resampled onto their original, native space by applying a single, composite transform to correct for head-motion and susceptibility distortions.

The BOLD time-series were resampled into the MNI152NLin2009cAsym standard space and used for further custom preprocessing. First, a binarized grey matter mask was generated by averaging the tissue probability maps of all subjects and thresholding at 95% grey matter probability. The grey matter mask and Gaussian smoothing (6mm kernel) were then applied to the preprocessed BOLD data. The first 10 samples, corresponding to the first 10 seconds of the task, were removed from all BOLD time series to minimize contributions of large stimulus onset responses (Nastase et al., 2019). The following confounds were regressed out of the BOLD signals: six translation and rotation parameters and their square, derivative, and squared derivative, and the CSF component output by fMRIPrep. Confound regression, high-pass filtering (140-second period), linear detrending, and z-scoring were applied using the nilearn signal.clean function in Python (Abraham et al., 2014).

Movie viewing quality assurance questionnaire

Following the verbal recall task, participants completed a quality assurance questionnaire to assess their recognition and comprehension of the movie, both scored from 0-1, in which they indicated whether they remembered seeing various items that may have appeared (e.g., airplane, gun, syringe) and answered yes/no questions about events that may have occurred (e.g., “Did the main character try to quit using drugs?”), and indicated whether they had seen the movie previously. Participants were also asked to rate, on a scale of 0-9, their attention to the movie, understanding of the narrative, and perception of the audio and video quality. Statistical comparisons between the groups are presented in Table S1 (Table S2 for the baseline replication sample). No significant group differences were observed at Session 1 in either the original or the replication sample (p≥.067); at Session 2, HUD participants scored lower on recognition (p=.028) and rated their understanding (p=.016) and the audio quality (p=.022) higher than CTL participants, although these effects were not significant after Bonferroni correction for the six separate tests.

Word selection for term categories of interest and control categories

Term categories and word lists are presented in Table S3. Categories of interest included first- (FP) and third-person (TP) pronouns and drug-related words. Pronoun word counts included all instances of singular and plural FP and TP terms. Since words related to drugs can be context-specific, these were selected manually by two independent raters from every unique word in the transcripts to include words that were used exclusively in reference to drugs and/or drug administration, acquisition, or abstinence, including incorrect/misspoken words (e.g., we determined that the words “suspicatory” and “repository” were intended to mean “suppository,” so these were included as drug words); words that were used in both drug and non-drug contexts in this dataset were not included.

To assess the specificity of the results to our term categories of interest, we also included two control categories (place words and stopwords) for which we did not expect to find group or session differences. Place words were selected from the transcripts through the same process as the drug words and included words used exclusively to refer to a location, including both general (e.g., “city”) and specific (e.g., “Edinburgh”) terms. Stopwords included commonly used terms that support the grammatical structure of a sentence but do not hold significant meaning; this list was derived from NLTK’s English stopword dictionary, accounting for lemmatization and expansion of contractions, with personal pronouns removed due to our specific interest in FP and TP pronouns. The mixed ANOVAs comparing control word frequencies by Group (CTL, HUD) and Session (1, 2) yielded no significant main or interaction effects (F_≤_2.14, p≥.148) (Fig. S1).

Baseline replication analyses

*Participant characteristics*: To assess the replicability of the language results observed at baseline, verbal recall samples from 32 additional HUD and 2 CTL participants who completed only Session 1 were analyzed along with the original 31 CTL participants (total 33 CTL/32 HUD). These included participants who withdrew from the study after Session 1, left the treatment facility, or were unable to complete the movie task procedures at Session 2. Twenty-nine of these HUD participants were taking methadone, and 3 buprenorphine/naloxone, as part of their inpatient treatment. The HUD replication sample did not differ significantly from the CTL group in age, gender, or nonverbal IQ, but did differ in race (p=.038), with more white and fewer Black and other participants in the HUD group; however, this effect was not significant after Bonferroni correction for the 4 separate demographic variables. The groups also differed in education and verbal IQ (CTL>HUD), depression and anxiety symptoms, and alcohol dependence (HUD>CTL). Similar to the original sample, all but one CTL participant reported never using nicotine regularly, and all HUD participants reported current or past regular nicotine use, so no statistical test was performed. Statistical tests for these variables and descriptive statistics for other drug use/addiction measures in the HUD group are presented in Table S2. For the analyses that involved fMRI data, three participants (1 CTL/2 HUD) were excluded due to a scanner malfunction that resulted in data loss but did not disrupt the movie viewing, and five HUD participants were excluded due to excessive head motion; as a result, 32 CTL/25 HUD were included in these analyses.

*Classification analysis*: Classifier performance on decoding the group labels (65 samples/2 classes) was assessed in the replication sample with the first 25 principal components of the embeddings, representing a cumulative variance ratio of 83.8%. Similar to the original sample, classification by group was significantly better than chance at Session 1 (true accuracy: 0.72, mean null accuracy: 0.50, p<.001) (Fig. S2).

*Semantic similarity analysis*: Within- (CTL-CTL, HUD-HUD) and between-groups (HUD-CTL) semantic similarity were assessed in the replication sample using the same procedures described in the main text. The 1-way ANOVA showed a significant main effect of Comparison (F_2,94_=8.76, p<.001); Tukey HSD post hoc testing revealed CTL-CTL>HUD-HUD (p=.002), CTL-CTL>HUD-CTL (p=.001), and HUD-HUD=HUD-CTL (p=.955) (Fig. S3).

*Drug and pronoun term frequencies analysis*: An independent groups t-test examining drug term frequencies in the replication sample showed a significant difference between the groups, with HUD>CTL (t_63_=-2.12, p=.038). For pronoun term frequencies, a 2-way ANOVA for Group (CTL, HUD) and Type (FP, TP) showed no main effects (F≤3.27, p≥.073), but a significant interaction effect of Group × Type (F_1,126_=19.15, p<.001), with independent groups t-tests revealing HUD>CTL for FP (t_63_=-4.05, p<.001) and CTL>HUD for TP (t_63_=2.25, p=.028). Paired t-tests comparing FP and TP frequencies showed no significant difference in HUD (t_31_=1.23, p=.228) and TP>FP in CTL (t_32_=-4.56, p<.001). These results are presented in Fig. S4.

Intersubject correlation analysis

To assess group and session effects on synchronization of participants’ fMRI signal during movie viewing, linear mixed effects modeling was performed on the network-level intersubject correlation scores (ISCs). Network-level fMRI signals were computed by averaging the z-scored BOLD time series data within eight major functionally defined networks from the Schaefer atlas 400 parcellations (Yeo et al., 2011). Network-level ISCs were computed using Pearson correlations between participants’ network-level time series data, which were then Fisher z-transformed for all subsequent analyses. These network ISCs are visualized by group and session in Fig. S6. A Group (HUD, CTL) × Session (1, 2) linear mixed effects model of network-level ISCs was performed with crossed random effects to account for non-independence between subject pairs (Chen et al., 2017). The p-values for main and interaction effects were FDR corrected across networks. Post-hoc comparison tests were performed via permutation testing, for which raw p-values are reported.

Results revealed main effects of Group (CTL>HUD) for default mode (β=-0.34, p<0.001), dorsal attention (β=-1.05, p<0.001), ventral attention (β=-0.17, p=0.02), somatomotor (β=-0.19, p=0.04), and visual (β=-0.99, p<0.001) networks, and main effects of Session (1>2) for dorsal attention (β=-0.17, p<0.001) and visual (β=-0.26, p<0.001) networks. There was a significant Group × Session interaction effect in default mode (β=0.19, p<0.001), dorsal attention (β=0.29, p<0.001), and visual (β=0.41, p<0.001) networks. Post hoc tests showed lower ISCs for HUD vs. CTL at Session 1 in all three networks (p<0.010), and at Session 2 for dorsal attention and visual networks (p<0.020). Within the HUD group, ISCs increased from Session 1 to Session 2 in all three networks (p<0.010); within the CTL group, ISCs decreased in dorsal attention and visual networks (p<0.002).

Intersubject representational similarity analysis

Results of the semantic-neural intersubject RSA, performed on the pairwise cosine similarities of the embeddings and intersubject correlations of the BOLD time series, in the combined networks are presented in Table S5 (Table S6 for the baseline replication sample). For this analysis, time series data from all ROIs within each of the major functionally defined networks in the Schaefer atlas were concatenated and assessed similarly to the ROI-based analysis, with Bonferroni correction of p-values for eight separate tests. Similar to the original sample, results from the replication sample showed significant HUD>CTL correlations in the dorsal attention and visual networks.

The ROI analysis in the baseline replication sample also yielded similar results to the original sample at Session 1, with HUD>CTL correlations in 21 cortical areas encompassing the bilateral dorsolateral prefrontal cortex/frontal eye field, superior parietal lobule, occipital fusiform gyri, and lateral occipital cortex; the left medial occipital cortex; and right occipital pole, as well as the bilateral temporal fusiform gyrus and the right supramarginal gyrus and lingual gyrus (significant results in the latter three were not found in the original sample) (Table S7 and Fig. S5). These areas similarly belonged to the dorsal attention and visual networks, except for the two supramarginal gyrus ROIs, which were labeled as temporoparietal regions.

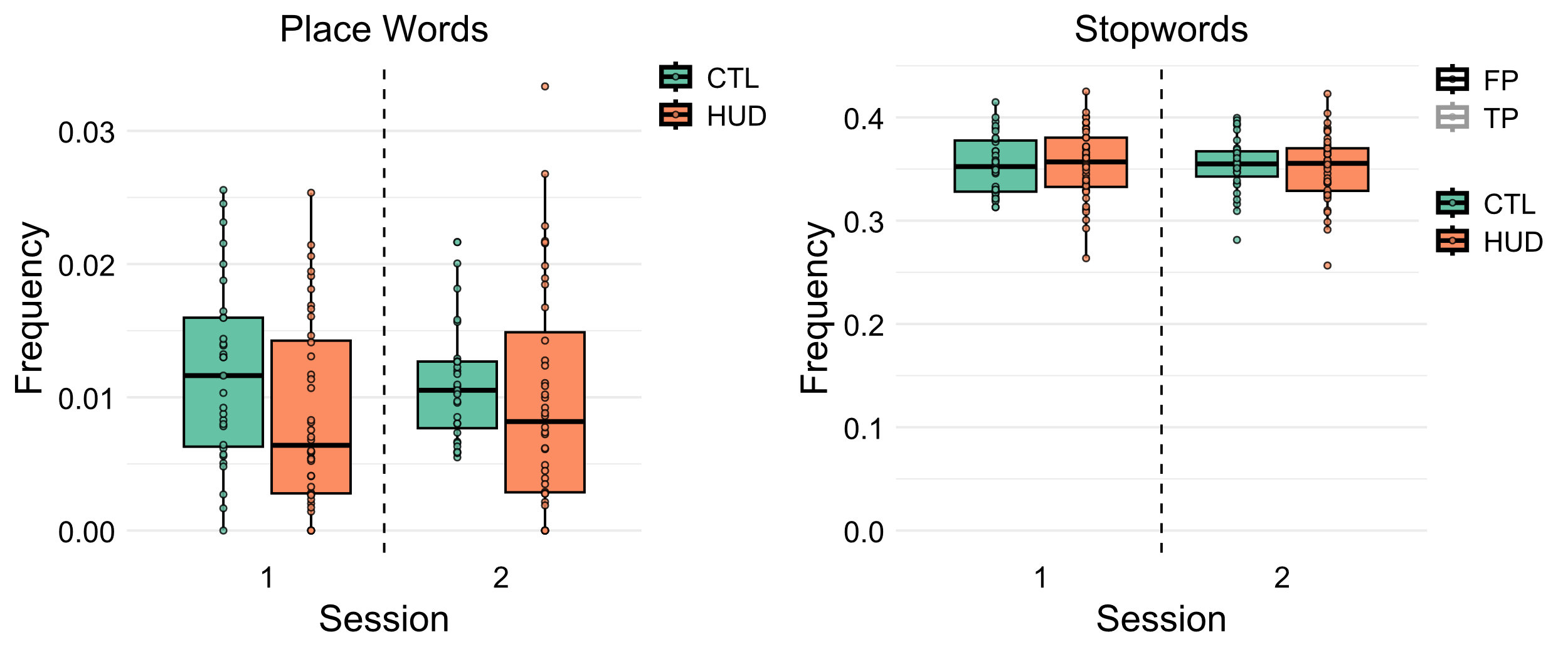

**Fig. S1. Term frequency analyses for control categories.** There were no significant differences in the frequency of place words or stop words between groups and sessions. In the boxplots, the center line indicates the median, the box indicates the interquartile range, and whiskers extend to the most extreme points within 1.5x the interquartile range above and below the box.

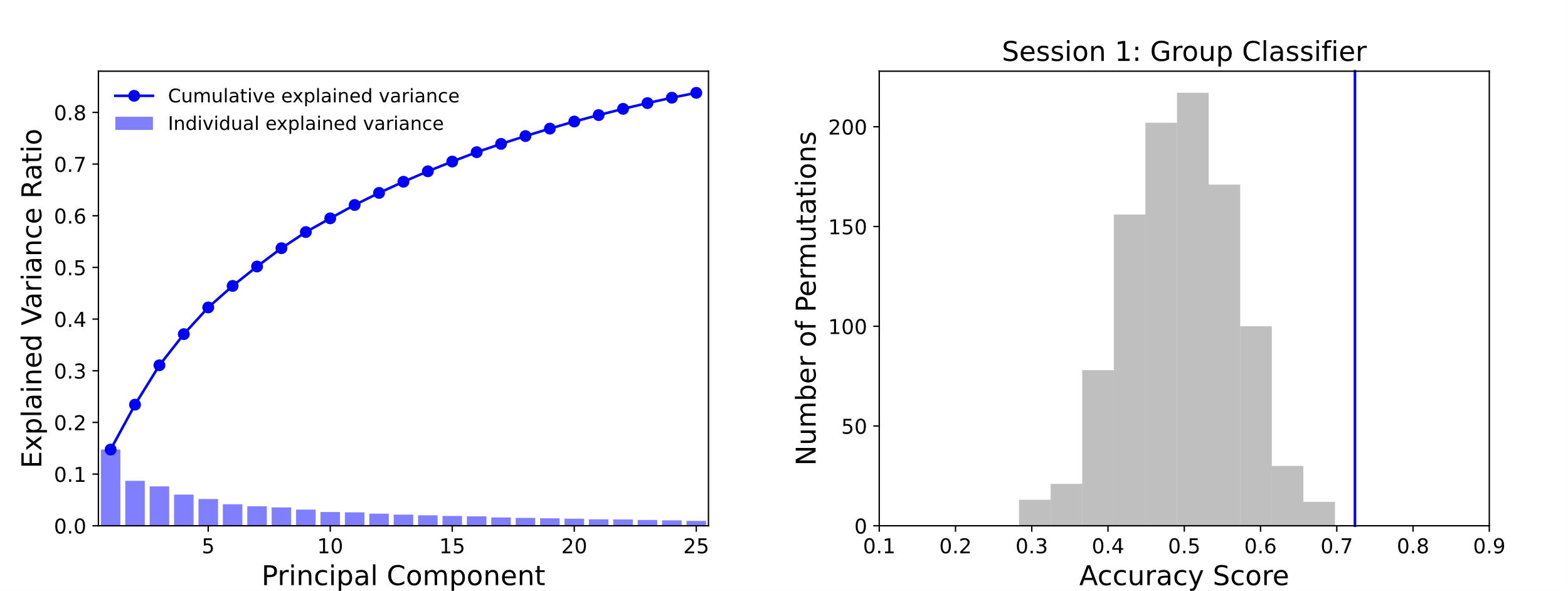

**Fig. S2 (related to Fig. 1). Classifier performance in the replication dataset at Session 1 on the first 25 principal components of the embeddings**. Principal components explained 83.8% of the total variance (variance plot) in the replication sample. In the classifier plot, the blue line indicates the true accuracy score, and the gray histogram represents the null accuracy distribution derived from 1,000 permutations of shuffled labels. Classification accuracy was significantly better than chance in decoding the group labels (0.72).

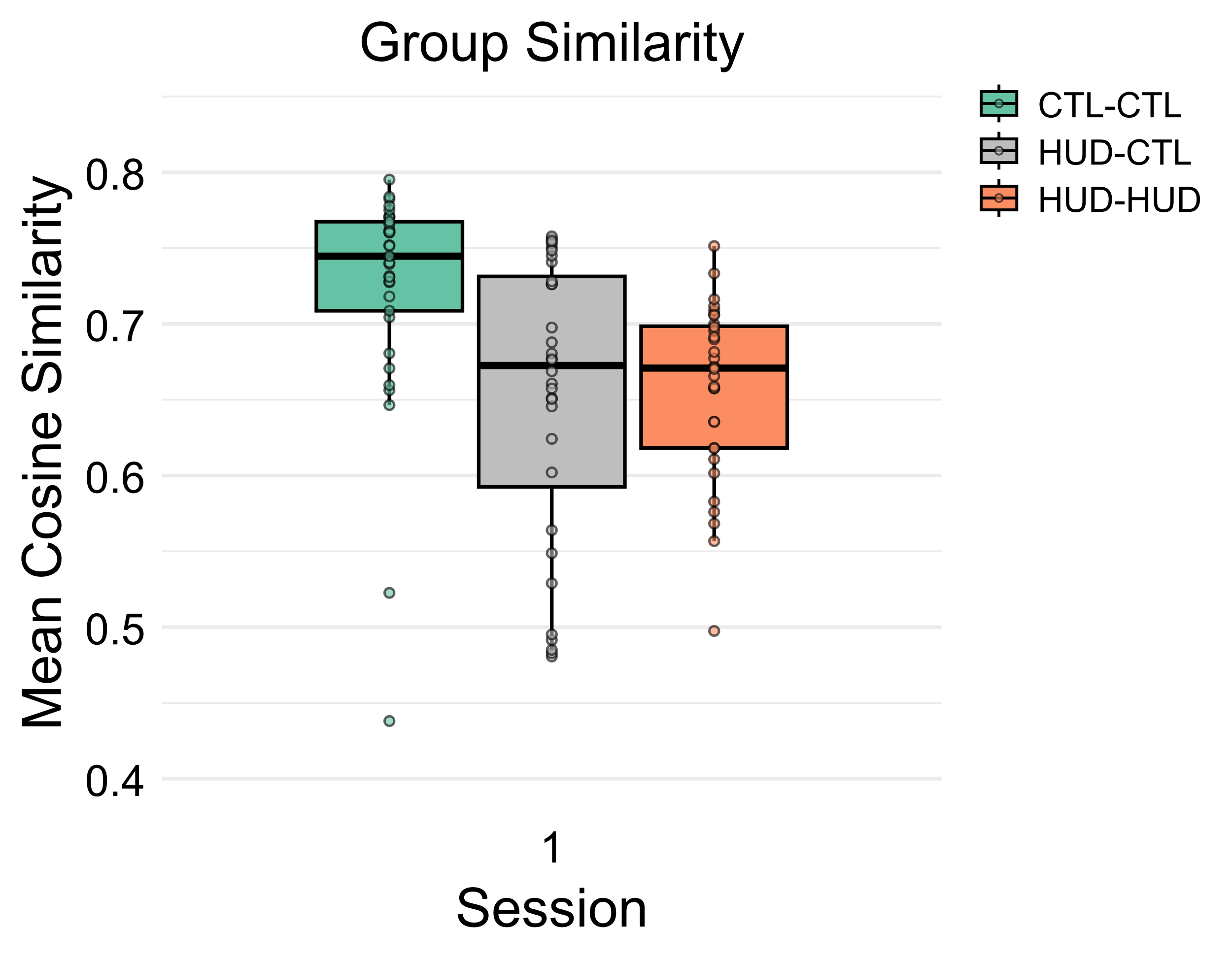

**Fig. S3 (related to Fig. 2B). Within- and between-group semantic similarities in the replication sample at Session 1.** Similar to the original dataset, the HUD group showed significantly lower within-group (HUD-HUD) similarity of embeddings than CTLs (CTL-CTL), with similar within- and between-group (HUD-CTL) similarity. In the boxplots, the center line indicates the median, the box indicates the interquartile range, and whiskers extend to the most extreme points within 1.5x the interquartile range above and below the box.

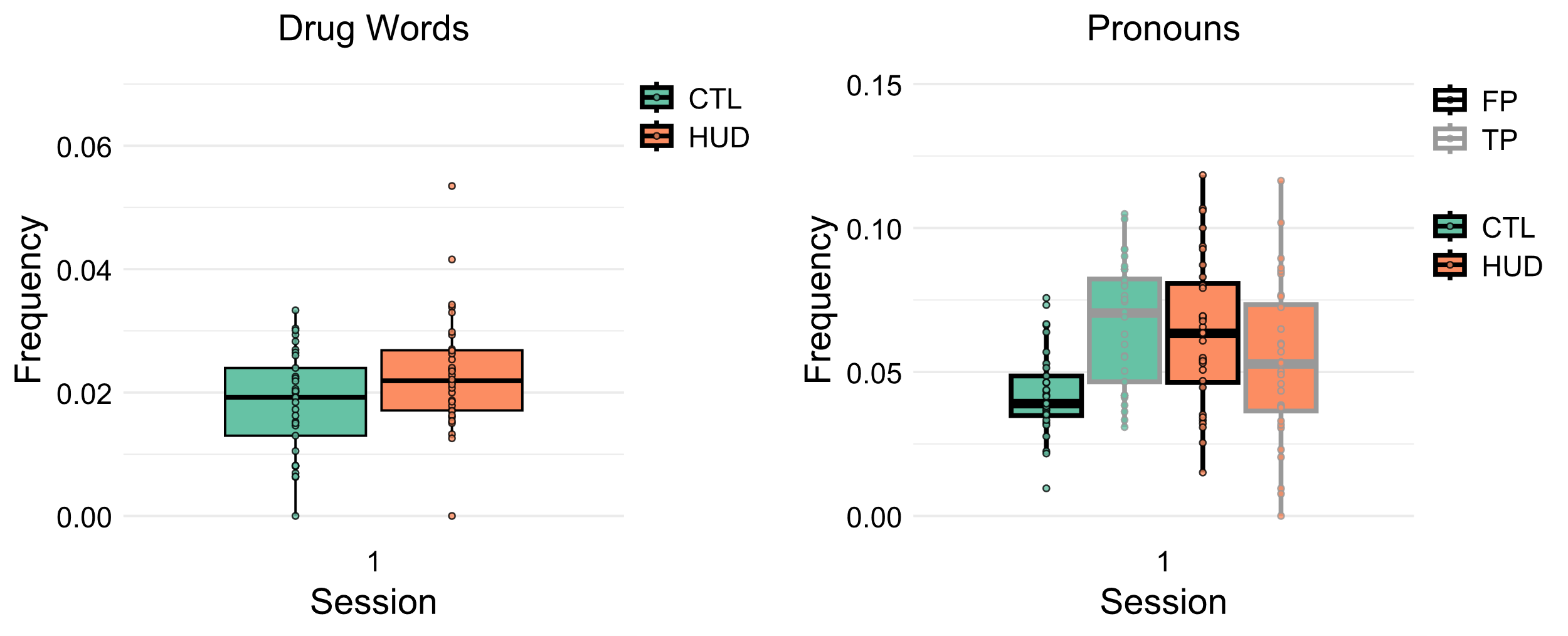

**Fig. S4 (related to Fig. 3). Term frequency analyses for categories of interest in the replication sample at Session 1.** Similar to the original dataset, the HUD group used drug words and FP pronouns at significantly higher frequencies, and TP pronouns and lower frequencies, than the CTL group. In the boxplots, the center line indicates the median, the box indicates the interquartile range, and whiskers extend to the most extreme points within 1.5x the interquartile range above and below the box.

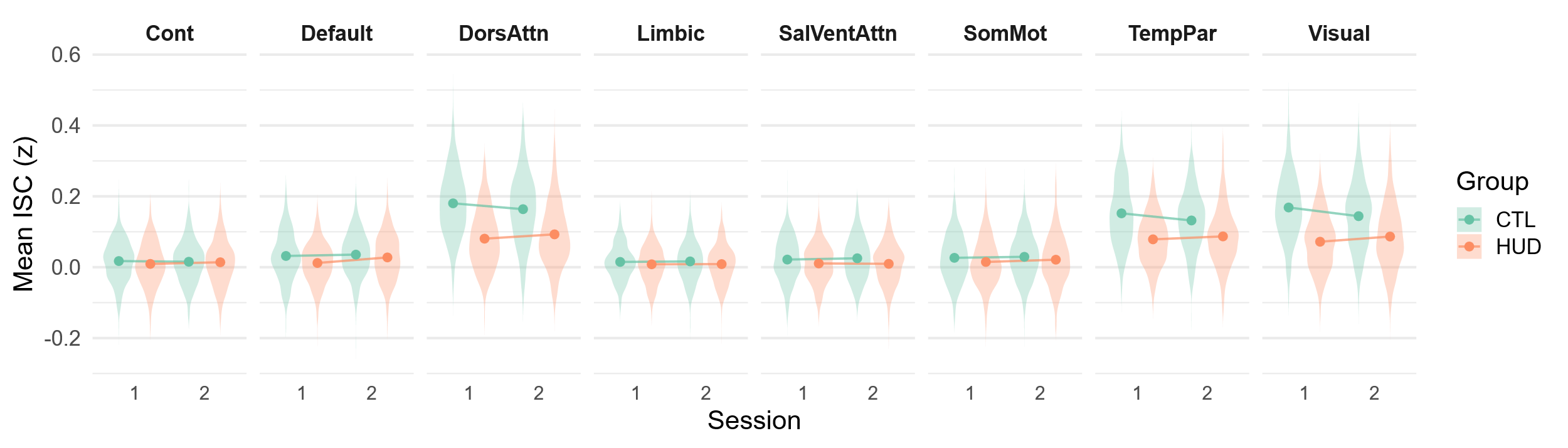

**Fig. S5. Mean Fisher z-transformed network ISCs by session and group.** Cont: Control; Default: Default Mode; DorsAttn: Dorsal Attention; SalVentAttn: Salience/Ventral Attention; SomMot: Somatomotor; TempPar: Temporoparietal.

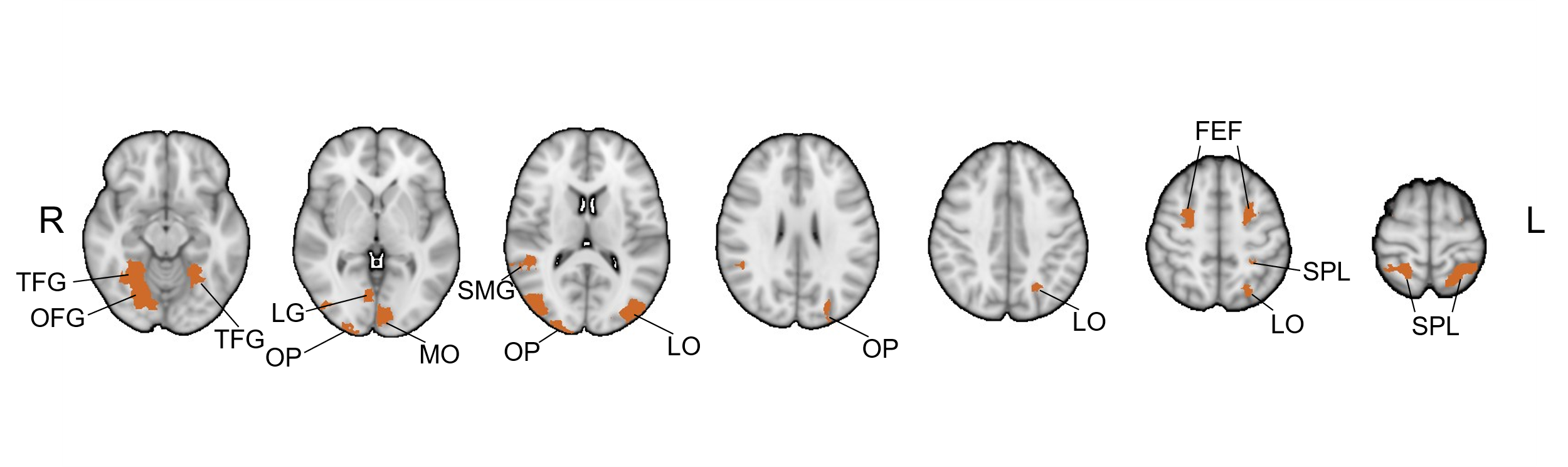

**Fig. S6 (related to Fig. 4). Anatomical distribution of the RSA results in the baseline replication sample.** Regions in which the Mantel test Spearman correlation was significantly greater in the HUD vs. CTL group, and which showed a significant positive correlation in the HUD group (see Table S3), are shown here. TFG: temporal fusiform gyrus; OFG: occipital fusiform gyrus; OP: occipital pole; LG: lingual gyrus; MO: medial occipital cortex; SMG: supramarginal gyrus; LO: lateral occipital cortex; FEF: frontal eye field; SPL: superior parietal lobule.

**Table S1 – Movie viewing quality assurance questionnaire**

|  | CTL, n=31 | HUD, n=40 | Test statistic | p-value |
| --- | --- | --- | --- | --- |
| Session 1 | | | | |
| **Recognition and comprehension** |  |  |  |  |
| Recognition, mean (SD) score | 0.73 (0.11) | 0.74 (0.11) | W=628 | .930 |
| Comprehension, mean (SD) score | 0.62 (0.13) | 0.63 (0.15) | W=594 | .744 |
| Seen previously, # yes/no | 21/10 | 22/18 | χ^2^=1.19 | .256 |
| **Movie quality** |  |  |  |  |
| Attention, mean (SD) rating | 6.8 (1.8) | 6.3 (2.2) | W=684 | .455 |
| Understanding, mean (SD) rating | 5.4 (2.7) | 6.3 (2.7) | W=473 | .085 |
| Audio quality, mean (SD) rating | 2.6 (2.2) | 3.5 (3.2) | W=554 | .442 |
| Video quality, mean (SD) rating | 6.1 (1.9) | 6.8 (2.3) | W=467 | .071 |
| Session 2 | | | | |
| **Recognition and comprehension** |  |  |  |  |
| Recognition, mean (SD) score | 0.76 (0.13) | 0.69 (0.13) | W=783.5 | .028 |
| Comprehension, mean (SD) score | 0.62 (0.15) | 0.62 (0.15) | W=612.5 | .871 |
| **Movie quality** |  |  |  |  |
| Attention, mean (SD) rating | 6.8 (2.1) | 6.8 (2.4) | W=583.5 | .841 |
| Understanding, mean (SD) rating | 5.4 (2.3) | 6.6 (2.7) | W=400 | .016 |
| Audio quality, mean (SD) rating | 3.1 (2.8) | 4.7 (3.0) | W=409 | .022 |
| Video quality, mean (SD) rating | 7.0 (1.9) | 7.1 (2.3) | W=535 | .429 |

**Notes**: Table presents mean and standard deviation (SD) or quantities for baseline sample characteristics. Test statistics were derived from two-sample Wilcoxon signed-rank test (W) for continuous variables and chi-squared test (χ^2^) for categorical variables. Missing data: 1 CTL participant Session 2 questionnaire.

**Table S2 – Movie viewing quality assurance questionnaire for the baseline replication sample**

|  | CTL, n=33 | HUD, n=32 | Test statistic | p value |
| --- | --- | --- | --- | --- |
| Session 1 | | | | |
| **Recognition and comprehension** |  |  |  |  |
| Recognition, mean (SD) score | 0.73 (0.12) | 0.70 (0.13) | W=611.5 | .273 |
| Comprehension, mean (SD) score | 0.61 (0.13) | 0.61 (0.13) | W=527 | .994 |
| Seen previously, # yes/no | 22/11 | 22/10 | χ^2^=0.03 | .857 |
| **Movie quality** |  |  |  |  |
| Attention, mean (SD) rating | 6.7 (1.8) | 5.9 (1.7) | W=666 | .067 |
| Understanding, mean (SD) rating | 5.3 (2.6) | 5.7 (2.1) | W=496.5 | .682 |
| Audio quality, mean (SD) rating | 2.6 (2.1) | 2.8 (2.3) | W=501.5 | .730 |
| Video quality, mean (SD) rating | 6.2 (1.9) | 6.4 (2.1) | W=484.5 | .567 |

**Notes**: Table presents mean and standard deviation (SD) or quantities for baseline sample characteristics. Test statistics were derived from two-sample Wilcoxon signed-rank test (W) for continuous variables and chi-squared test (χ^2^) for categorical variables.

| **Table S3 - Term categories and word lists** | |
| --- | --- |
| **Category** | **Words** |
| drug | addict, addicted, addiction, administer, administering, alcohol, alcoholic, aspirin, baggie, baggies, balloon, bar, beer, boffed, boffing, boof, boofed, boofing, boosted, boosting, boozed, boozing, bundle, cap, caplets, capsule, carrier, cigarette, cocaine, coke, cooker, cooking, copped, copping, crack, craves, craving, dab, dealer, den, deposit, depository, detox, dope, dopesick, dose, drug, drug-taking, drug-wise, drugging, drunk, feening, fiended, fix, free-basing, gallery, habit, heroin, high, highness, hooked, illicit, inebriated, inhale, inject, injected, injecting, injection, injects, intravenously, iv, joint, junkie, junky, keister, loony, lsd, marijuana, medication, medicine, meth, methadone, moonie, narcotic, needle, od, opiate, opioid, opium, over-the-counter, overdosed, overdosing, pack, packet, paraphernalia, pill, pill-wise, pipe, pouch, powder, powdered, powdery, prescription, psychedelics, puff, quit, recreational, repository, shoot-up, shooter, slapping, smacked, smoke, smoking, sniffed, sniffer, sniffing, snorting, sober, speed, spoon, stash, stashed, stimulant, strapped, strung, substance, suppository, suspicatory, syringe, syrinx, tablet, tapping, tobacco, tourniquet, trap, turkey, tying, usage, user, valium, vein, wasted, weed, whiskey, wine, withdrawal |
| first-person | i, me, mine, my, myself, our, ours, ourselves, us, we |
| third-person | he, her, hers, herself, him, himself, his, she, their, theirs, them, themselves, they |
| place | airport, alcove, america, apartment, area, balcony, bar, barn, basement, bathroom, beach, bedroom, bridge, britain, budapest, building, chicago, chinatown, city, club, corner, country, den, diner, doorway, downtown, dwelling, edinburgh, england, environment, establishment, europe, field, gallery, ghetto, hall, hallway, hill, home, hotel, house, ireland, island, jail, kitchen, lawn, loft, london, nation, neighborhood, nigeria, nightclub, office, park, parlor, pathway, penn, philly, portland, portugal, property, restaurant, restroom, rikers, road, school, scotland, shop, sidewalk, staircase, stall, station, street, theater, toronto, town, trap, tunnel, uk, underground, undersea, underwater, united, upstairs, warehouse, yard |
| stopwords | what which who whom this that these those am is are was were be been being have has had having do does did doing a an the and but if or because as until while of at by for with about against between into through during before after above below to from up down in out on off over under again further then once here there when where why how all any both each few more most other some such no nor not only own same so than too very s t can will just don should now |

**Table S4 (related to Table 1) – Baseline demographic, neuropsychological, and drug use/addiction measures for the baseline replication sample**

|  | CTL, n=33 | HUD, n=32 | Test statistic | p-value |
| --- | --- | --- | --- | --- |
| **Demographics** |  |  |  |  |
| Age, mean (SD) years | 40.5 (11.0) | 39.8 (9.1) | W=546.5 | .911 |
| Gender, # men/women | 22/11 | 27/5 | χ^2^=2.75 | .098 |
| Race, # Black/white/multiracial or other | 10/15/8 | 3/24/5 | χ^2^=6.52 | .038 |
| Education, mean (SD) years | 16.1 (3.0) | 11.6 (2.3) | W=899 | <.001 |
| **Neuropsychological tests** |  |  |  |  |
| WRAT – mean (SD) Reading standard score | 109.2 (6.6) | 96.9 (11.4) | W=858 | <.001 |
| WASI – mean (SD) Matrix Reasoning scaled score | 11.8 (2.7) | 10.5 (3.0) | W=656 | .091 |
| Depression symptoms, mean (SD) BDI score | 4.4 (9.4) | 13.2 (10.8) | W=201.5 | < .001 |
| Anxiety symptoms, mean (SD) BAI score | 3.2 (6.5) | 11.5 (11.8) | W=261 | <.001 |
| **Drug use and addiction variables** |  |  |  |  |
| Regular nicotine use, # current/past/never^a^ | 1/5/27 | 29/3/0 | - | - |
| Alcohol dependence, mean (SD) SMAST score | 0.3 (0.9) | 3.3 (3.7) | W=183 | <.001 |
| Heroin use disorder |  |  |  |  |
| Age of onset, mean (SD) years | - | 22.7 (5.4) | - | - |
| Regular use, mean (SD) years | - | 12.4 (7.0) | - | - |
| Current abstinence, mean (SD) days | - | 194.2 (196.5) | - | - |
| Past 30-day use, mean (SD) days | - | 0.9 (5.3) | - | - |
| Craving symptoms, mean (SD) HCQ score | - | 43.9 (15.9) | - | - |
| Withdrawal symptoms, mean (SD) SOWS score | - | 3.8 (5.4) | - | - |
| Severity of dependence, mean (SD) SDS score | - | 12.2 (3.2) | - | - |

**Notes**: Table presents mean and standard deviation (SD) or quantities for baseline sample characteristics for the replication sample. Test statistics were derived from two-sample Wilcoxon signed-rank test (W) for continuous variables and chi-squared test (χ^2^) for categorical variables. ^a^Since only one CTL participant reported current nicotine use and only two HUD participants reported non-current nicotine use, no statistical test was performed on this measure. Missing data: 1 CTL and 1 HUD datapoint for education; 2 HUD datapoints for BDI and BAI; 2 datapoints for regular heroin use; 1 datapoint for HCQ and SOWS.

**Table S5 – Statistics for network-level semantic-neural representational similarity analysis**

|  | Correlation Comparison, Z_diff_ (p_corr_) | | | | Session 1, r_s_ (p) | | Session 2, r_s_ (p) | |
| --- | --- | --- | --- | --- | --- | --- | --- | --- |
| Schaefer Network Label | Group: Session 1 | Group: Session 2 | Session: HUD | Session: CTL | CTL | HUD | CTL | HUD |
| Control | 1.45 (>.999) | 0.88 (>.999) | 2.66 (.062) | - | 0.07 (.256) | **0.16 (.023)** | -0.05 (.683) | <0.01 (.467) |
| Default mode | 1.31 (>.999) | 0.71 (>.999) | 1.32 (>.999) | - | 0.07 (.244) | **0.16 (.017)** | 0.04 (.348) | 0.08 (.197) |
| Dorsal attention | **3.67 (.002)** | 2.70 (.055) | 2.42 (.125) | - | -0.01 (.528) | **0.21 (.025)** | -0.09 (.746) | 0.08 (.250) |
| Limbic | 0.94 (>.999) | -0.22 (>.999) | 1.65 (.794) | - | 0.06 (.189) | **0.12 (.011)** | 0.03 (.297) | 0.02 (.368) |
| Salience/Ventral attention | -1.36 (>.999) | 0.63 (>.999) | 0.60 (>.999) | - | 0.17 (.058) | 0.09 (.087) | 0.01 (.427) | 0.05 (.231) |
| Somatomotor | -0.47 (>.999) | 0.54 (>.999) | 0.51 (>.999) | - | 0.10 (.169) | 0.07 (.129) | 0.01 (.444) | 0.04 (.273) |
| Temporoparietal | -2.05 (0.32) | -0.66 (>.999) | 0.27 (>.999) | - | 0.17 (.095) | 0.04 (.316) | 0.07 (.275) | 0.03 (.393) |
| Visual | **4.12 (<.001)** | 2.52 (.093) | **2.83 (.038)** | - | -0.04 (.585) | **0.22 (.027)** | -0.10 (.760) | 0.06 (.297) |

**Notes**: Table presents statistics for networks evaluated in representational similarity analysis of recall semantic similarity and BOLD ISC during movie viewing in the replication sample. Comparisons of the correlation coefficients between groups/within each session, and between sessions within HUD, were performed using Fisher Z testing (Z_diff_) with Bonferroni-corrected p values, shown in the Correlation Comparison column. Spearman r (r_s_) and uncorrected p-values derived from Mantel tests are shown for each group. Bold values indicate statistically significant tests.

**Table S6 – Statistics for network-level semantic-neural representational similarity analysis in the baseline replication sample**

|  | Correlation Comparison, Z_diff_ (p_corr_) | Session 1, r_s_ (p) | |
| --- | --- | --- | --- |
| Schaefer Network Label | Group: Session 1 | CTL | HUD |
| Control | 1.62 (.835) | 0.07 (.237) | **0.19 (.038)** |
| Default mode | 2.44 (.117) | 0.08 (.234) | 0.25 (.252) |
| Dorsal attention | **4.16 (<.001)** | 0.03 (.404) | **0.32 (.008)** |
| Limbic | -0.48 (>.999) | 0.05 (.198) | 0.02 (.389) |
| Salience/Ventral attention | 0.53 (>.999) | **0.17 (.046)** | **0.21 (.011)** |
| Somatomotor | 0.06 (>.999) | 0.11 (.125) | 0.11 (.104) |
| Temporoparietal | 1.87 (.489) | 0.19 (.070) | **0.32 (.007)** |
| Visual | **4.27 (<.001)** | 0.03 (.414) | **0.33 (.009)** |

**Notes**: Table presents statistics for networks evaluated in representational similarity analysis of recall semantic similarity and BOLD ISC during movie viewing in the replication sample. Comparisons of the correlation coefficients between groups were performed using Fisher Z testing (Z_diff_) with Bonferroni-corrected p values, shown in the Correlation Comparison column. Spearman r (r_s_) and uncorrected p-values derived from Mantel tests are shown for each group. Bold values indicate statistically significant tests.

**Table S7 (related to Table 3) – Statistics for semantic-neural representational similarity analysis for the baseline replication sample**

|  | Correlation Comparison, Z_diff_ (p_corr_) | Session 1, r_s_ (p) | |
| --- | --- | --- | --- |
| Structure (Schaefer Atlas #) | Group: Session 1 | CTL | HUD |
| **HUD>CTL** |  |  |  |
| Left Lateral Occipital Cortex (10)^1^ | **5.22 (<.001)** | -0.08 (.723) | **0.29 (.020)** |
| Left Lateral Occipital Cortex (12)^1^ | **4.23 (.010)** | 0.01 (.475) | **0.31 (.019)** |
| Left Temporal Fusiform Gyrus (13)^1^ | **4.17 (.013)** | 0.12 (.202) | **0.40 (.002)** |
| Left Medial Occipital Cortex (18)^1^ | **4.48 (.003)** | 0.03 (.378) | **0.34 (.003)** |
| Left Temporal Fusiform Gyrus (61)^1^ | **3.95 (.032)** | 0.08 (.204) | **0.36 (<.001)** |
| Left Superior Parietal Lobule (68)^1^ | **4.12 (.016)** | -0.01 (.510) | **0.28 (.008)** |
| Left Superior Parietal Lobule (78)^1^ | **3.93 (.035)** | -0.12 (.841) | **0.16 (.043)** |
| Left Superior Parietal Lobule (80)^1^ | **4.99 (<.001)** | -0.12 (.825) | **0.24 (.029)** |
| Left Frontal Eye Field (83)^1^ | **4.40 (.004)** | -0.01 (.502) | **0.31 (<.001)** |
| Right Temporal Fusiform Gyrus (201)^1^ | **4.32 (.006)** | 0.07 (.284) | **0.37 (<.001)** |
| Right Occipital Fusiform Gyrus (203)^1^ | **5.30 (<.001)** | -0.01 (.508) | **0.37 (.007)** |
| Right Occipital Pole (209)^1^ | **3.86 (.047)** | -0.07 (.734) | **0.21 (.049)** |
| Right Lateral Occipital Cortex (210)^1^ | **5.65 (<.001)** | -0.04 (.596) | **0.36 (.006)** |
| Right Occipital Pole (211) | **3.95 (.033)** | -0.09 (.758) | 0.19 (.067) |
| Right Lateral Occipital Cortex (212) | **4.04 (.012)** | -0.07 (.677) | 0.22 (.062) |
| Right Temporal Fusiform Gyrus (213)^1^ | **4.79 (<.001)** | 0.05 (.352) | **0.38 (.003)** |
| Right Lingual Gyrus (215)^1^ | **4.19 (.012)** | -0.02 (.541) | **0.28 (.012)** |
| Right Superior Parietal Lobule (280)^1^ | **5.11 (<.001)** | -0.07 (.692) | **0.29 (.003)** |
| Right Frontal Eye Field (283)^1^ | **3.99 (.028)** | 0.03 (.385) | **0.31 (.002)** |
| Right Supramarginal Gyrus (397)^1^ | **4.14 (.015)** | 0.03 (.346) | **0.33 (.008)** |
| Right Supramarginal Gyrus (399)^1^ | **4.61 (.002)** | 0.04 (.362) | **0.36 (.001)** |

**Notes**: Table presents statistics for structures that met criteria for significance in representational similarity analysis of recall semantic similarity and BOLD ISC during movie viewing in the replication sample. Comparisons of the correlation coefficients between groups were performed using Fisher Z testing (Z_diff_) with Bonferroni-corrected p values, shown in the Correlation Comparison column. Spearman r (r_s_) and uncorrected p-values derived from Mantel tests are shown for each group. Bold values indicate statistically significant tests (p<.05). Regions that showed a significant group difference in the correlation coefficient and significant RSA correlation within HUD are denoted ^1^.
